# Regional Vulnerability to Brain Metastases Reflects Structural, Metabolic and Genetic Organization of the Healthy Brain

**DOI:** 10.64898/2026.11.06.26367469

**Authors:** Joseph I. Turner, Ariel Arias, Ross MacFadyen, Allen Y. Fu, Raymond Y. Huang, Jay Patel, Nayan Lamba, Hesham Elhalawani, Ayal A. Aizer, Aaron Yannis Schwieger, Fabian Wolpert, Marian Galovic, Michael Weller, Kenneth Bernstein, Michael E. Pacold, Eric K. Oermann, Frederic L. W. V. J. Schaper, Douglas Kondziolka

## Abstract

**Background:** Brain metastases (BMs), the most common intracranial tumor, do not distribute uniformly throughout the brain, but instead preferentially localize to specific regions. Building on known associations with perfusion and the gray-white matter junction, we hypothesized that regional vulnerability to BMs also reflects the cellular and molecular organization of the healthy brain.

**Methods:** In a discovery cohort of 1,406 patients, we generated a voxelwise map of BM distribution and compared it with normative atlases of brain microstructure, vasculature, metabolism, cell-type composition, and gene expression. Associations were tested for replication in an independent multicenter validation cohort of 1,209 patients.

**Results:** In the discovery cohort, BM frequency was higher near the gray-white matter junction and in regions with greater cerebral perfusion, glucose metabolism, and mitochondrial density. At the cellular level, BM frequency was inversely related to astrocyte and microglial transcriptional signatures. Using normative gene expression maps derived from postmortem healthy brain tissue, we identified 17 genes spatially associated with BM vulnerability and specific to metastases relative to other intracranial lesion types. These associations replicated in the validation cohort. The immune checkpoint gene *LAG3* ranked among the ten genes most positively associated with metastatic vulnerability across all genes tested (*r* = 0.40-0.45, *p* < 0.001). *LAG3* expression was also elevated in peritumoral brain metastasis tissue relative to control brain tissue in an independent spatial transcriptomic dataset.

**Conclusions:** These findings suggest that BM vulnerability reflects healthy brain organization and identify LAG-3, a clinically targetable immune checkpoint, as a candidate link between regional brain immune biology and metastatic colonization.

## INTRODUCTION

Brain metastases (BMs) are the most common intracranial tumor, occurring in 10-26% of adults with systemic cancer and outnumbering primary brain malignancies roughly ten to one.^1,2^ Patients with BMs carry a poor prognosis,^3,4^ with survival substantially worse than that conferred by systemic disease alone.^5^ Successful colonization of the brain requires cancer cells to adapt to a microenvironment unlike that of any other organ. The brain is separated from the circulation by the blood-brain barrier and contains specialized resident cells, such as astrocytes and microglia, that are not found elsewhere in the body. BMs are therefore increasingly understood as a distinct central nervous system disease rather than a simple extension of the primary tumor.^6^

BMs are not uniformly distributed throughout the brain but show a well-documented predilection for posterior and infratentorial structures.^7–10^ This pattern is consistent with the “seed and soil” hypothesis, in which regional tissue properties act as permissive “soil” for metastatic “seeds.”^11,12^ Prior work has linked this regional vulnerability to cerebral perfusion and proximity to the gray-white matter junction.^13,14^

Here, we tested whether regional vulnerability also reflects the cellular and molecular organization of the healthy brain. Large, publicly available lesion datasets and normative brain atlases now make it possible to test this across the whole brain. Using our previously developed open-source atlas of brain metastasis vulnerability^15^ (n = 1,406) as a Discovery cohort and an independent multicenter Validation cohort (n = 1,209), we compared metastasis frequency maps with normative atlases of brain physiology, cell-type organization, and gene expression.

## METHODS

### Patient Cohorts and Brain Metastasis Atlas Construction

The Discovery cohort comprised 1,406 patients with brain metastases treated with Gamma Knife radiosurgery at NYU Langone Health (NYUMets dataset).^16,17^ Baseline T1-weighted MRI scans and radiosurgery target contours were nonlinearly normalized to the MNI152 template using EasyReg, a deep learning deformable registration algorithm in FreeSurfer,^18^ and superimposed to generate a voxelwise metastasis frequency map.^15^ The Validation cohort comprised 1,209 patients pooled from three independent cohorts: Brigham and Women’s Hospital and Dana-Farber Cancer Institute (BWH/DFCI; n = 702; segmented via algorithm with manual correction),^19,20^ the University of Zurich (n = 193; manual expert segmentations),^21^ and UCSF (n = 314; manual expert segmentation).^22^ All Validation lesions were normalized to MNI152 space using EasyReg,^18^ summed voxelwise across datasets, and divided by the combined sample size so each patient contributed equally. For both cohorts, we also constructed etiology-stratified atlases for lung, breast, melanoma, and other primary malignancies. Patient characteristics and lesion metrics are summarized in Supplementary Table S1; cohort inclusion and segmentation protocols are detailed in the Supplementary Methods.

### Normative Brain Atlases

We assembled 14 normative volumetric atlases in MNI152 space spanning microstructural organization (synaptic density,^23^ myelin,^24^ gray and white matter tissue probability,^25,26^ and gray-white matter junction probability calculated as the voxelwise product of gray and white matter probabilities), vascular architecture (cerebral blood flow,^27^ cerebral blood volume,^28^ medium-to-large arterial density,^29^ vessel radius,^30,31^ and venous density, diameter, and torsion^32,33^), and metabolism (mitochondrial density^34^ and glucose metabolism^35^). Sources are listed in Supplementary Table S2.

Whole-brain transcriptomic maps for 15,633 genes were obtained from the Allen Human Brain Atlas (AHBA)^36^ using previously generated voxelwise expression maps^37^ (N. Nolan, unpublished manuscript) processed with the abagen toolbox^38^ via standard filtering, hemispheric mirroring, probe selection, and donor normalization parameters (Supplementary Methods). To estimate regional cell-type signatures across the 31 cell-type superclusters defined in the Human Brain Cell Atlas,^39^ we extracted the AHBA spatial expression maps for each supercluster’s canonical marker genes (median 23 genes per cell type) and computed their first principal component across voxels to capture shared spatial variance (Supplementary Methods).

### Spatial Correlation and Statistical Analysis

All normative atlases and metastasis frequency maps were resampled to 2 mm isotropic resolution, and spatial associations were quantified as voxelwise Pearson correlations (Fig. S1). Significance was assessed against 5,000 null maps in which lesions were randomly relocated within the brain parenchyma while preserving lesion count and volume (Supplementary Methods).^15^ Two-sided empirical *P* values were corrected with the Benjamini-Hochberg false discovery rate (FDR) procedure within each biological category (microstructural, vascular, metabolic, cellular, and transcriptomic; α = 0.05). Features significant in the Discovery cohort were tested in the Validation cohort, with FDR correction across those features, and were considered reproducible if significant in both cohorts with concordant direction. Analyses were performed in Python and R (packages and versions in Supplementary Methods).

### Transcriptomic, Checkpoint, and Sensitivity Analyses

Candidate genes were nominated in three steps. First, from the full set of 15,633 gene expression maps, we selected the 10 genes most positively and 10 most negatively correlated with the Discovery metastasis map. Second, we retained genes whose correlation with the Validation metastasis map fell in the same tail of the distribution of Validation correlations across all 15,633 genes (two-tailed α = 0.025, meaning above the 98.75th or below the 1.25th percentile). Third, the surviving candidates (17 of 20) were re-tested with the spatial permutation framework, with FDR correction across candidates; all 17 met FDR-corrected *P* < 0.001 in both the Discovery and Validation cohorts. Disease specificity was assessed by comparing normative expression averaged within patient-level metastasis masks against public lesion masks of stroke,^40^ meningioma,^41^ focal cortical dysplasia,^42^ and multiple sclerosis,^43,44^ and matched random lesions, using FDR-corrected Mann-Whitney U tests. Independence from physiological features was assessed by re-correlating gene maps against the residuals of an ordinary least squares model predicting Discovery metastasis frequency from the reproducible microstructural, vascular, and metabolic features. Cross-cohort gene rank reproducibility was assessed with Spearman’s rank correlation. Pathway enrichment was evaluated on ranked Discovery correlation lists using gene set enrichment analysis (GSEA)^45^ across Gene Ontology Biological Processes and cell-type marker sets^39^ (FDR α = 0.05), and significant sets were re-tested in the Validation cohort (Supplementary Methods).

To verify that candidate associations were not driven solely by cerebellar-cerebral differences, spatial correlations and 5,000-permutation tests were repeated restricting all maps to a supratentorial mask (Supplementary Methods). Reproducible physiological, cellular, and transcriptomic associations were also recalculated across etiology-stratified atlases, including melanoma-specific candidate nomination and GSEA (Supplementary Methods). Finally, we compared six canonical immune checkpoint genes (*PDCD1*, *CD274*, *CTLA4*, *LAG3*, *HAVCR2*, and *TIGIT*)^46^ with the Discovery and Validation maps as well as a map combining both cohorts for a single summary of spatial similarity; Fig. 6A), and confirmed *LAG3* expression across human non-small cell lung cancer brain metastasis spatial RNA-sequencing data^47^ (comparing peritumoral [n = 20] vs. control [n = 7] samples via Welch’s t-test), healthy pig brain regions,^48^ and single-cell human brain cells^39^ (Supplementary Methods).

## RESULTS

The Discovery cohort comprised 1,406 patients treated with Gamma Knife radiosurgery at NYU Langone Health, whose lesion locations we have described previously.^15^ The Validation cohort comprised 1,209 patients pooled from three previously described cohorts at Brigham and Women’s Hospital and Dana-Farber Cancer Institute (BWH/DFCI),^19^ UCSF,^22^ and the University of Zurich^21^ (Supplementary Table S1). Discovery cohort lesions were the tumor target contours drawn for radiosurgery planning. Validation cohort lesions were segmented manually by experts (UCSF, Zurich) or by a segmentation algorithm with manual review and correction (BWH/DFCI, see Methods). In each cohort, lesions were spatially normalized to a common anatomical template and superimposed to generate a voxelwise map of metastasis frequency (Fig. 1A). Consistent with prior reports,^8,14^ metastasis frequency varied across brain regions in both cohorts.

**Figure 1.**
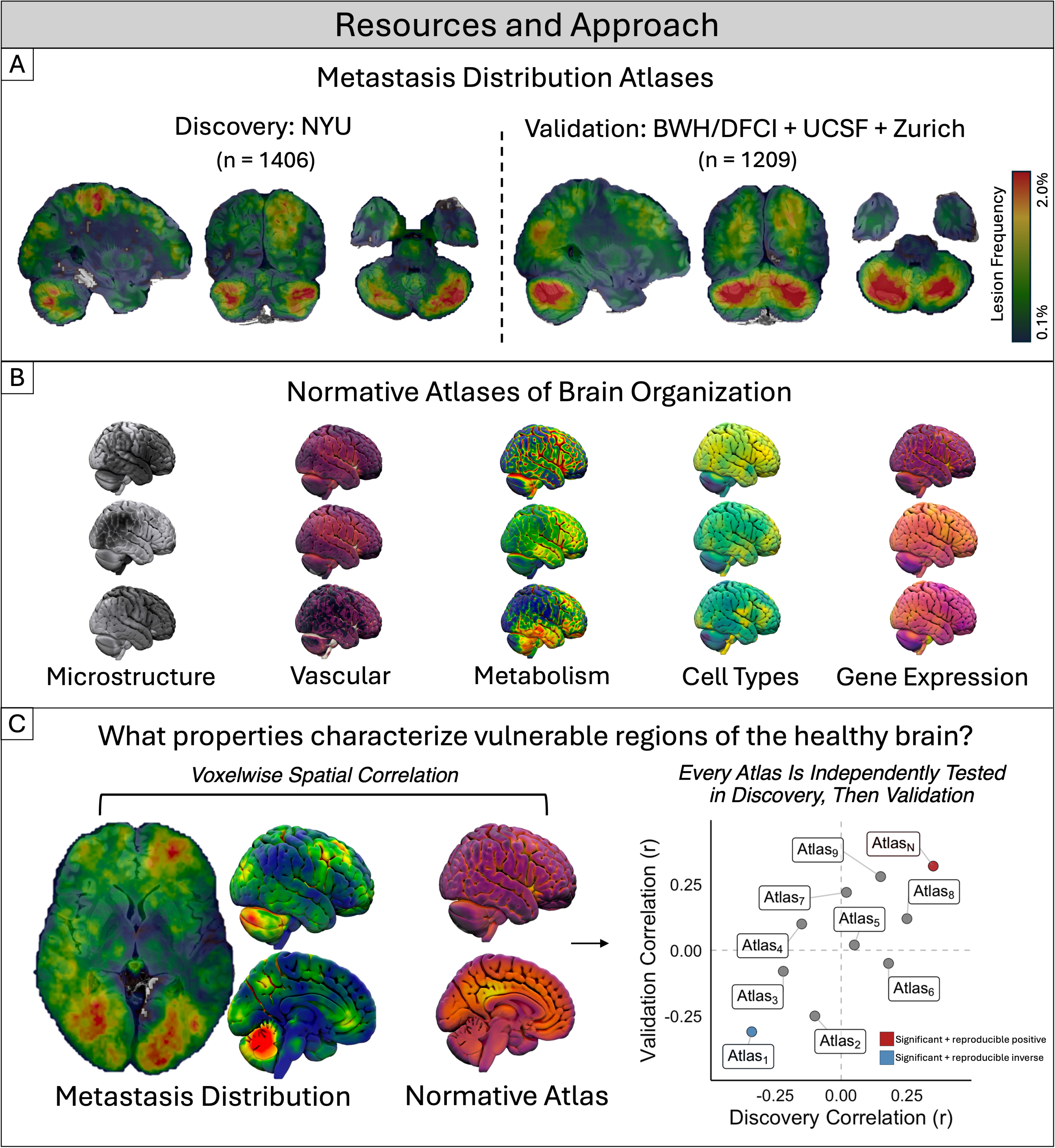
Resources and approach for characterizing spatial patterns of brain metastasis vulnerability. **(A)** Voxelwise metastasis distribution atlases generated by spatially normalizing all lesions to MNI152 space in the Discovery (NYU, n = 1,406) and Validation (Brigham and Women’s Hospital and Dana-Farber Cancer Institute, UCSF, and University of Zurich; n = 1,209) cohorts. Warmer colors indicate higher lesion frequency across the cohort. **(B)** Normative atlases of healthy brain organization spanning microstructural, vascular, metabolic, cell-type, and transcriptomic properties, each derived independently of the metastasis data. **(C)** Schematic of the spatial correlation approach. Metastasis distribution maps were spatially correlated on a voxelwise basis with each normative atlas independently in the Discovery and Validation cohorts. Associations were considered reproducible only if statistically significant in both cohorts after correction for multiple comparisons and concordant in direction of effect. Right, each point represents one normative atlas positioned according to its Discovery and Validation correlation coefficients. Red and blue indicate atlases with significant, reproducible positive and inverse associations, respectively.

To test whether regional properties of brain tissue act as the “soil” in the “seed and soil” model of metastasis,^11,12^ we assembled a panel of normative atlases spanning microstructural, vascular, metabolic, cell-type, and transcriptomic brain organization, each derived independently of any metastasis data (Fig. 1B; Supplementary Table S2). We spatially correlated each atlas with the metastasis frequency maps on a voxelwise basis (Fig. 1C). Significance was assessed by spatial permutation testing, in which lesion locations were randomized throughout the brain parenchyma while preserving the number and volume of observed lesions. Associations were identified in the Discovery cohort and tested for replication in the Validation cohort; only associations that remained significant after multiple-comparison correction in both cohorts, with concordant directionality, were considered reproducible.

### Metastases preferentially localize to metabolically active, highly perfused regions at the gray-white junction

Consistent with prior studies,^13,14^ BMs localized preferentially to the gray-white matter junction and to highly perfused regions, and additionally favored metabolically active regions supplied by smaller-caliber vessels (Fig. 2; all FDR-corrected *P* < 0.05 within category). Of five microstructural atlases, only gray-white junction probability was reproducibly associated with BM frequency (Discovery *r* = 0.20, Validation *r* = 0.12). Of seven vascular atlases, three were reproducibly associated: cerebral blood flow positively (Discovery *r* = 0.24, Validation *r* = 0.15), and vessel radius (Discovery *r* = -0.22, Validation *r* = -0.28) and intermediate-sized vessel density (Discovery *r* = -0.19, Validation *r* = -0.22) inversely. Both metabolic atlases were positively associated, including mitochondrial density (Discovery *r* = 0.20, Validation *r* = 0.21) and glucose metabolism (Discovery *r* = 0.28, Validation *r* = 0.20). Because these MRI-derived vascular atlases resolve only medium-to-large arteries, the inverse associations indicate that BMs occur less often where large vessels predominate, not that metastasis-prone regions are less vascularized overall. Together with the positive association with cerebral blood flow, this pattern suggests that BMs favor well-perfused tissue supplied predominantly by smaller vessels.

**Figure 2.**
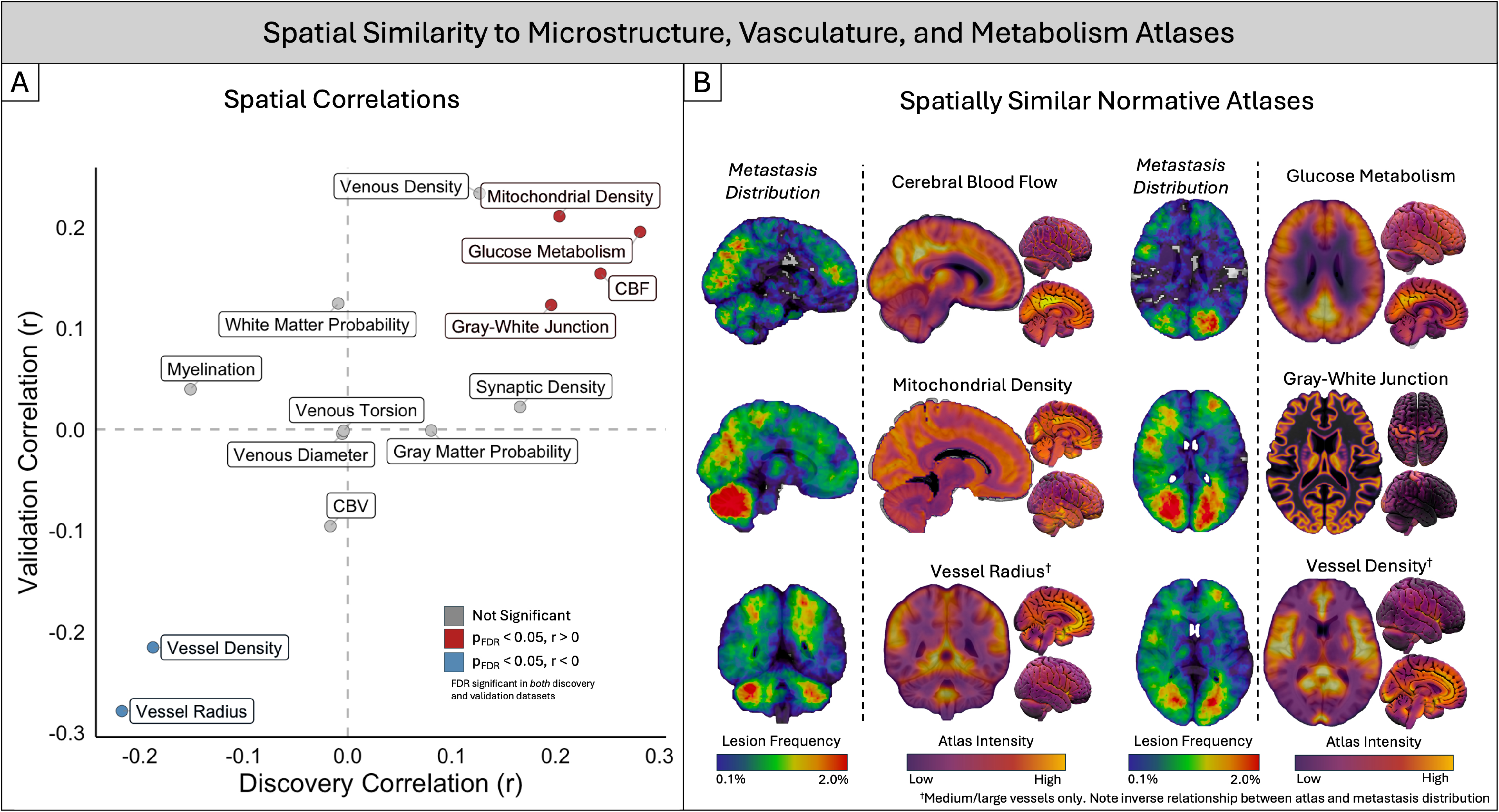
Brain metastases preferentially localize to metabolically active, highly perfused brain regions at the gray-white junction. **(A)** Spatial correlation between metastasis distribution and normative atlases of brain microstructure, vasculature, and metabolism, shown for the Discovery (x-axis) and Validation (y-axis) cohorts. Red and blue indicate atlases with statistically significant (FDR-corrected P < 0.05), reproducible positive and inverse spatial correlations, respectively, in both cohorts. Gray indicates atlases not meeting this threshold. **(B)** Representative spatial comparisons between metastasis distribution and each significantly and reproducibly associated normative atlas. † indicates an inverse spatial relationship to metastasis distribution (higher atlas intensity corresponds to lower lesion frequency, and vice versa). Vessel Radius and Vessel Density atlases capture only medium-to-large arterial structures resolvable by MRI and do not reflect microvascular or capillary-level density.

### Metastasis-prone regions show low astrocyte and microglial transcriptional signatures

To test whether metastasis distribution relates to the cellular composition of brain tissue, we compared the BM distribution maps to spatial maps of specific brain cell types. Because direct measurements of cell-type abundance are not available across the whole human brain at voxel resolution, we estimated cell-type maps from normative gene expression. For each cell-type supercluster defined in the Human Brain Cell Atlas,^39^ we took the marker genes identified by single-cell sequencing and summarized their spatial expression in the Allen Human Brain Atlas as the first principal component (Fig. 3). We focused primarily on non-neuronal cell types given their established roles in vascular regulation, immune signaling, and maintenance of the brain microenvironment. Of nine non-neuronal cell types, five showed reproducible associations: BMs occurred more frequently in regions with lower astrocyte (Discovery *r* = -0.39, Validation *r* = - 0.42), microglia (Discovery *r* = -0.36, Validation *r* = -0.36), oligodendrocyte precursor cell (Discovery *r* = -0.32, Validation *r* = -0.37), and fibroblast (Discovery *r* = -0.27, Validation *r* = - 0.28) signatures, and in regions with higher Bergmann glia signatures (Discovery *r* = 0.23, Validation *r* = 0.35) (Fig. 4; all FDR-corrected *P* < 0.05). Of 22 neuronal cell types, 12 showed reproducible associations, all inverse (Fig. S2).

**Figure 3.**
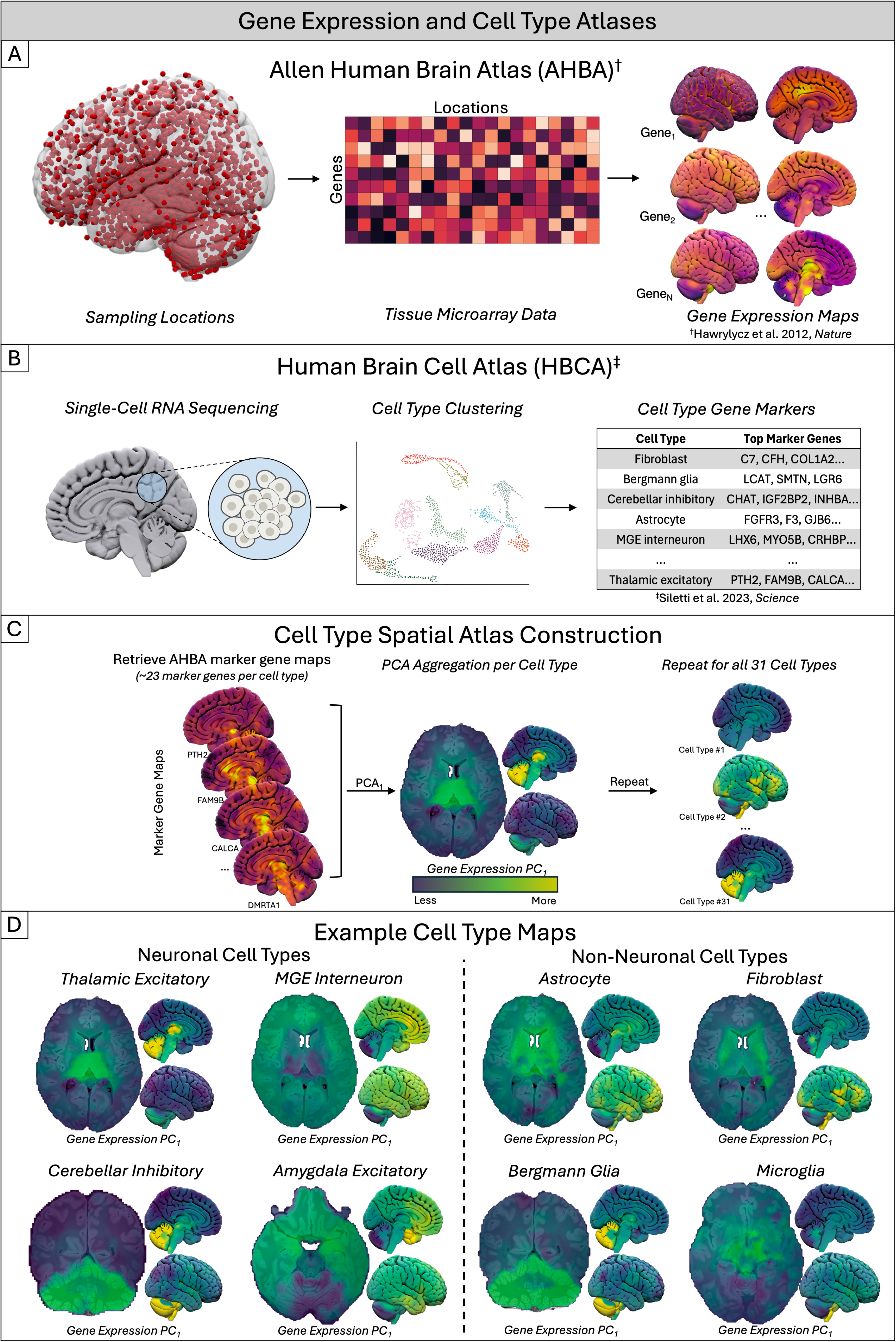
Construction of gene expression and cell-type spatial atlases. **(A)** The Allen Human Brain Atlas (AHBA) provides microarray-based gene expression measurements sampled from anatomically distributed tissue locations across the brain, yielding spatial expression maps for each gene.^†^ **(B)** The Human Brain Cell Atlas (HBCA) provides single-cell RNA sequencing data organized into 31 major cell-type superclusters, each defined by a characteristic set of marker genes.^‡^ **(C)** For each cell-type supercluster, spatial expression maps of its marker genes were retrieved from the AHBA, and the first principal component (PC1) of these marker-gene maps was computed to generate a single representative spatial atlas for that cell type. This process was repeated independently for all 31 cell-type superclusters. **(D)** Representative spatial atlases for four neuronal (thalamic excitatory, MGE interneuron, cerebellar inhibitory, amygdala excitatory) and four non-neuronal (astrocyte, fibroblast, Bergmann glia, microglia) cell-type superclusters, illustrating the distinct spatial organization captured by each cell-type atlas. Cell icon: NIAID Visual & Medical Arts. (10/7/2024). Generic Cells. NIAID NIH BIOART Source. bioart.niaid.nih.gov/bioart/172; UMAP Icon: SingleCell_Clustering_DataReduction_UMAP icon by James-Lloyd https://www.badgrammargoodsyntax.com/ is licensed under CC0 https://creativecommons.org/publicdomain/zero/1.0/

**Figure 4.**
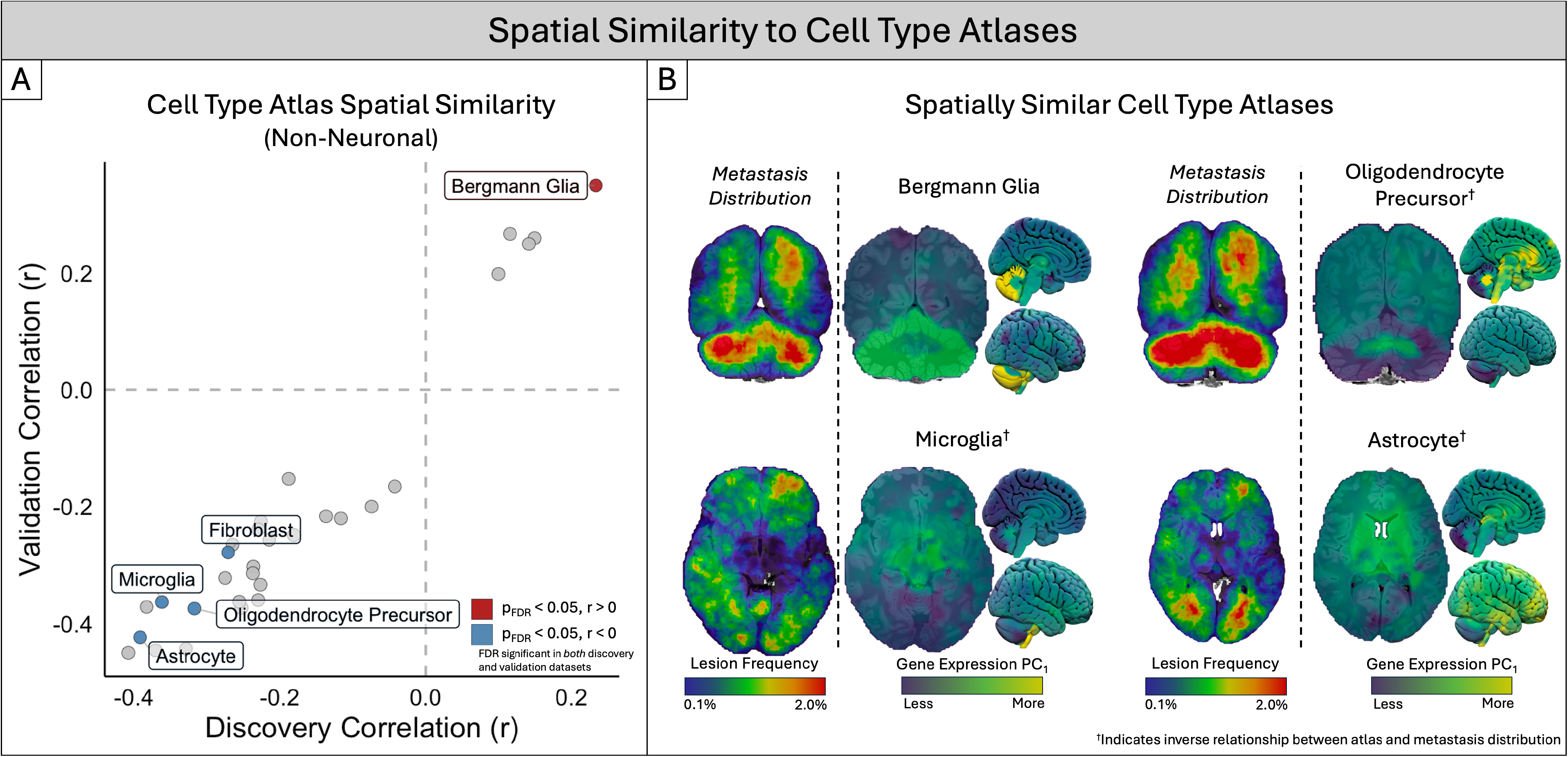
Brain metastases preferentially localize to regions with distinct cell-type identities. **(A)** Spatial correlation between metastasis distribution and cell-type atlases derived from the Human Brain Cell Atlas, shown for all 31 cell-type superclusters in the Discovery (x-axis) and Validation (y-axis) cohorts. Red and blue indicate cell types with statistically significant (FDR-corrected P < 0.05), reproducible positive and inverse spatial correlations, respectively, in both cohorts; gray indicates cell types not meeting this threshold. Only non-neuronal cell types, which are more directly relevant to metastatic seeding and the perivascular niche, are labeled; cell-type correlations with neuronal superclusters is provided in Fig. S2. **(B)** Representative spatial comparisons between metastasis distribution and each significantly and reproducibly associated non-neuronal cell-type atlas. † indicates cell types with an inverse spatial relationship to metastasis distribution.

Bergmann glia are found primarily in the cerebellum, where they perform astrocyte-like functions;^49^ their positive association may therefore largely reflect the high frequency of BMs in the cerebellum. Outside this cerebellum-specific population, BMs were more common in regions with weaker astrocyte and microglial signatures, suggesting that regional differences in glial composition relate to metastatic susceptibility.

### Metastasis-prone regions show high normative expression of *LAG3*, *FGF9*, and *ALDH1A3*

We next asked whether the spatial distribution of BMs resembles the expression pattern of specific genes in the healthy brain. The Allen Human Brain Atlas measured gene expression at thousands of sites in postmortem brains from six healthy donors, from which whole-brain maps of where each gene is expressed can be generated.^36,38^ We correlated the BM frequency maps with these maps for 15,633 genes. To nominate candidates, we required each gene to rank among the ten most positively or negatively correlated genes in the Discovery cohort and to replicate in the Validation cohort (see Methods). Seventeen genes (nine positive, eight negative) met both criteria (Fig. 5). These genes remained highly ranked after accounting for the gray-white junction, vascular, and metabolic features identified above (Fig. S3), indicating that their associations are not explained by regional blood flow or metabolism.

**Figure 5.**
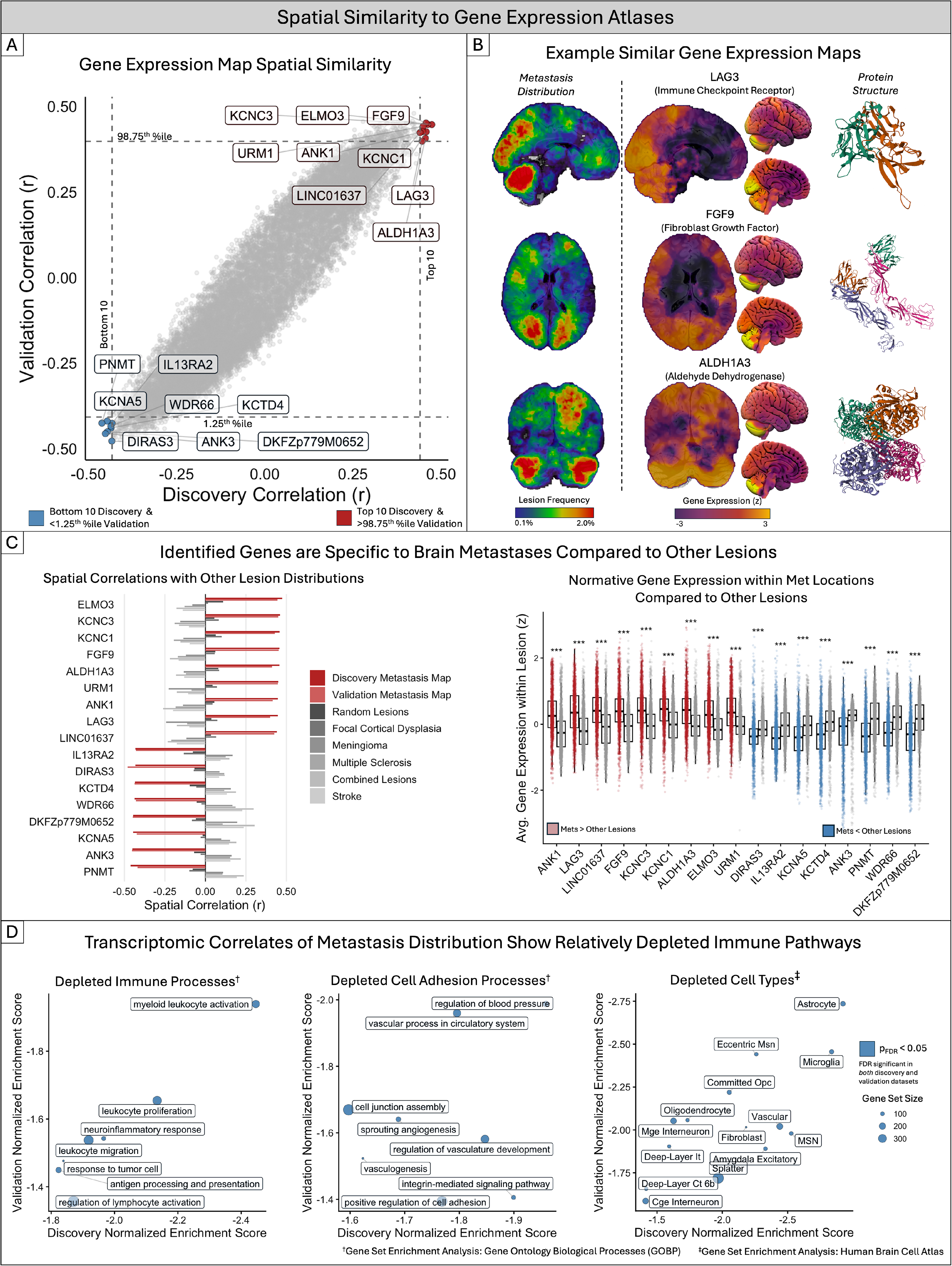
Spatial expression of individual genes identifies transcriptomic correlates of metastasis vulnerability and depleted immune pathways. **(A)** Spatial correlation between metastasis distribution and 15,633 gene expression maps from the Allen Human Brain Atlas, shown for the Discovery (x-axis) and Validation (y-axis) cohorts. Genes were considered candidate correlates if they ranked among the top or bottom 10 discovery correlations and additionally exceeded a two-tailed significance threshold in validation (α = 0.025 two tailed, corresponding to the 98.75th and 1.25th percentiles); nine positively and eight negatively associated genes met these criteria. **(B)** Representative spatial comparisons between metastasis distribution and three genes of particular biological interest among the top-ranked candidates (*LAG3*, *FGF9*, and *ALDH1A3*), with corresponding protein structures. **(C)** Left, spatial correlation between candidate gene expression maps and lesion distributions for brain metastases (Discovery and Validation) versus other intracranial pathologies, including randomly generated lesions, focal cortical dysplasia, meningioma, multiple sclerosis, and stroke (representative lesion distribution maps shown in Fig. S4). Right, normative expression of each candidate gene within metastasis lesion locations compared to other lesion types (***FDR-corrected P < 0.001). **(D)** Gene set enrichment analysis (GSEA) performed on the complete ranked list of gene-metastasis spatial correlations, rather than on the individual candidate genes highlighted in (A). GSEA tests whether genes belonging to predefined biological pathways or cell-type signatures are concentrated near the top or bottom of the ranked gene list, indicating coordinated enrichment or depletion across the transcriptome. Shown here are Gene Ontology biological processes^†^ and Human Brain Cell Atlas cell-type transcriptional programs^‡^ that were reproducibly depleted in metastasis-prone regions across Discovery and Validation cohorts. Point size indicates gene set size; blue indicates FDR-significant reproducibility in both cohorts. Only gene sets surviving FDR-correction independently in both cohorts are shown. LAG3 Structure : 10.2210/pdb7TZG/pdb; FGF9 structure: 10.2210/pdb5W59/pdb; ALDH1A3 structure: 10.2210/pdb5FHZ/pdb

Gene rankings across all genes were highly reproducible between cohorts (Spearman’s rho = 0.94). Among genes with higher expression in metastasis-prone regions, we highlight three representing distinct biological processes: *LAG3*, associated with immune regulation (Discovery *r* = 0.45, Validation *r* = 0.40); *FGF9*, associated with cellular differentiation (Discovery *r* = 0.46, Validation *r* = 0.45); and *ALDH1A3*, associated with metabolism (Discovery *r* = 0.46, Validation *r* = 0.41) (Fig. 5B). Genes with lower expression in metastasis-prone regions included *IL13RA2* (Discovery *r* = -0.43, Validation *r* = -0.43), whose expression was highest in regions relatively spared by metastases.

To test whether these associations were specific to BMs rather than a feature of any brain lesion, we compared candidate gene expression at metastasis locations with expression at the locations of other intracranial lesions, including focal cortical dysplasia, meningioma, multiple sclerosis, stroke, and randomly generated lesions (Fig. S4). Expression of all seventeen candidate genes differed significantly at metastasis locations relative to these other lesion types (all FDR-corrected *P* < 0.001, Fig. 5C).

To test whether broader groups of genes showed the same pattern, we performed gene set enrichment analysis, which asks whether genes sharing a biological function are concentrated at the top or bottom of the ranked list of gene-metastasis correlations. We tested Gene Ontology biological processes and Human Brain Cell Atlas cell-type transcriptional programs. Gene sets related to immune and glial function were reproducibly concentrated among genes with lower expression in metastasis-prone regions, including myeloid leukocyte activation, leukocyte proliferation, antigen presentation, and astrocyte and microglia transcriptional programs (Fig. 5D).

### Candidate associations persist within the supratentorial compartment

BMs occur disproportionately in the posterior fossa,^7–10^ and the cerebellum differs from the rest of the brain in its cellular and transcriptional programs.^50^ The associations above could therefore reflect features that distinguish the cerebellum from the cerebrum rather than variation within the cerebrum. To test this, we repeated the analysis using supratentorial voxels only (Fig. S5). Gene associations were modestly attenuated in the Discovery cohort (*LAG3*, *r* = 0.45 whole brain vs 0.41 supratentorial; *FGF9*, 0.46 vs 0.40; *ALDH1A3*, 0.46 vs 0.42; *IL13RA2*, -0.43 vs -0.38), and all 17 candidate genes remained significant in both cohorts by spatial permutation testing (all FDR-corrected *P* < 0.001). Within the supratentorial compartment, all cell-type associations also remained significant in both cohorts except Bergmann glia, a population restricted to the cerebellum. These results suggest that the candidate features partly reflect differences between the cerebellum and cerebrum, which may contribute to the posterior fossa predilection of BMs, but also track metastatic vulnerability outside the posterior fossa.

### The immune checkpoint gene *LAG3* is elevated in metastasis-vulnerable brain regions

We next examined six canonical immune checkpoint genes (*PDCD1* [PD-1], *CD274* [PD-L1], *CTLA4*, *LAG3*, *HAVCR2* [TIM-3], and *TIGIT*). *LAG3* showed the strongest positive spatial correlation with metastasis vulnerability (Fig. 6A; see Fig. S6 for correlations with the Discovery and Validation maps individually) and was also among the seventeen candidate genes identified across all genes above. LAG-3 is an inhibitory immune checkpoint receptor that suppresses T-cell activation and contributes to the immunosuppressed microenvironment in cancer.^51^ Gene set enrichment analysis also identified lower expression of immune-related gene sets in metastasis-prone regions (Fig. 5D), suggesting that immunoregulatory pathways may contribute to regional metastatic susceptibility.

**Figure 6.**
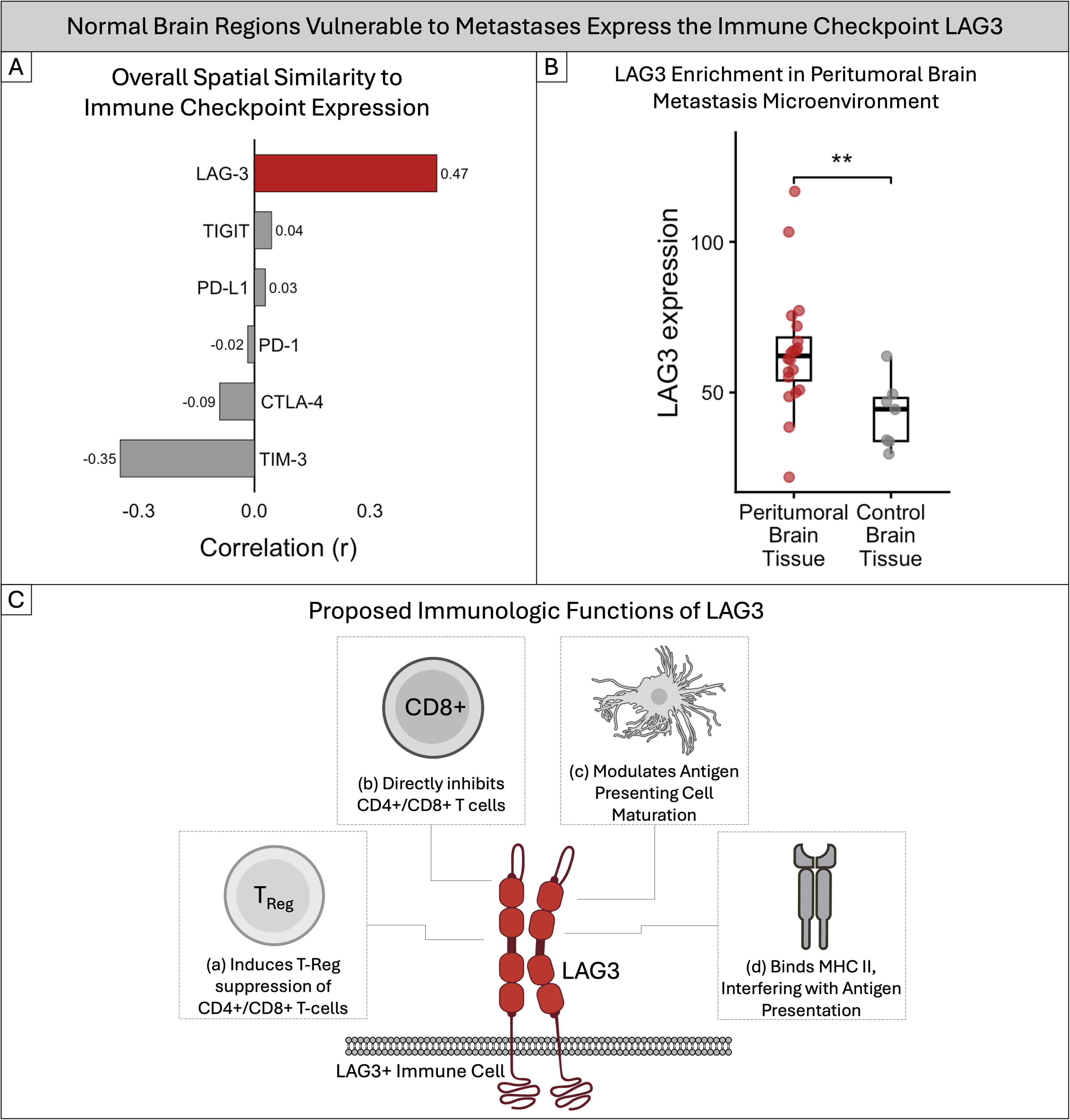
Normal brain regions vulnerable to metastases express the immune checkpoint LAG3. **(A)** Spatial correlation between the combined Discovery and Validation metastasis distribution map and normative expression of canonical immune checkpoint genes (*PDCD1* [PD-1], *CD274* [PD-L1], *CTLA4*, *LAG3*, *HAVCR2* [TIM-3], and *TIGIT*). Among the checkpoint genes examined, *LAG3* showed the strongest spatial association with metastasis vulnerability. Correlations with the individual Discovery and Validation maps are provided in Fig. S6. **(B)** *LAG3* expression in peritumoral tissue (n = 20) compared with control brain tissue (n = 7) from surgically resected non-small cell lung cancer brain metastases (Welch’s *t*-test; *P* < 0.01).^47^ **(C)** Proposed immunologic functions of LAG-3 relevant to the metastatic niche. LAG-3 has been reported to act through several non-mutually exclusive mechanisms: (i) LAG-3 on regulatory T cells (Tregs) enhances Treg-mediated suppression of CD4+ and CD8+ effector T cells; (ii) LAG-3 on CD4+ and CD8+ T cells inhibits their activation and effector function upon ligand engagement; (iii) LAG-3 on Tregs modulates antigen-presenting cell maturation through contact-dependent interaction with MHC class II; and (iv) LAG-3 binds MHC class II, influencing antigen presentation. LAG3 icon inspired by 10.1038/s41590-023-01569-z; T cell icon: NIAID Visual & Medical Arts. (10/7/2024). T Cell. NIAID NIH BIOART Source. bioart.niaid.nih.gov/bioart/509; Dendritic cell icon: NIAID Visual & Medical Arts. (10/7/2024). Dendritic Cell. NIAID NIH BIOART Source. bioart.niaid.nih.gov/bioart/112; MHC Class II icon: NIAID Visual & Medical Arts. (10/7/2024). MHC Class II. NIAID NIH BIOART Source. bioart.niaid.nih.gov/bioart/343; Plasma membrane icon: 10.5281/zenodo.13690056

Because these findings relied on normative atlases derived from healthy brain tissue, we next tested whether *LAG3* expression is also elevated in the metastasis microenvironment. In an independent spatially resolved dataset of surgically resected human brain metastases and control brain tissue,^47^ *LAG3* expression was higher in peritumoral metastasis tissue than in control tissue (*t* = 3.28, *P* < 0.01) (Fig. 6B), suggesting that the normative association extends to diseased brain tissue.

As a further check, we confirmed that *LAG3* is expressed in healthy brain tissue in two additional datasets. In the healthy pig brain,^48^ *LAG3* expression was detectable across regions and peaked in the cerebellum. In a single-cell atlas of the healthy human brain,^39^ *LAG3* was expressed in T cells and other immune populations, as well as in populations not traditionally considered immune-related, such as Purkinje cells (Fig. S7, Supplementary Table S3).

### The metastasis vulnerability niche generalizes across carcinoma subtypes but diverges in melanoma

To assess whether the vulnerability niche generalizes across metastasis etiologies, we recalculated all significant, reproducible associations separately for etiology-stratified distributions of primary lung, breast, other carcinomas, and melanoma. Physiological, cell-type, and gene-level associations were broadly consistent across lung, breast, and other carcinomas. Melanoma diverged: cell-type correlations were consistently attenuated or reversed, and candidate genes, including *LAG3*, showed weaker or divergent associations (Fig. S8); gene-level reproducibility between Discovery and Validation was also lower for melanoma than for the pooled analysis (Spearman’s rho = 0.81; Fig. S9).

Applying the same candidate criteria to the melanoma-specific maps identified five reproducible genes: *TNNT2* and *PART1* were positively associated with melanoma metastasis frequency, and *MAF*, *HLA-DOA*, and *LINC00886* were inversely associated (Fig. S9B). Gene set enrichment analysis of melanoma-specific correlations further identified reproducible enrichment for gene sets involved in axon guidance and synaptic signaling in both cohorts (Fig. S9C). Leading-edge genes of these sets included components of Wnt and Eph/ephrin signaling, such as *WNT7A*, *EPHB2*, and *DVL1*, pathways implicated in melanoma invasion and metastatic growth.^52–54^ Overall, the features associated with brain metastasis vulnerability were largely shared across carcinoma metastases but differed for melanoma, a malignancy of neuroectodermal origin.

## DISCUSSION

Across two independent cohorts, brain regions frequently colonized by metastases aligned with vascular, metabolic, cellular, and molecular features of the healthy brain. Guided by the seed- and-soil hypothesis,^11^ we asked whether regional properties of the “soil” contribute to where metastases form, comparing metastasis distributions with normative atlases acquired independently of disease. Metastasis-prone regions showed higher cerebral perfusion, increased metabolic activity, greater proximity to the gray-white matter junction, lower astrocyte and microglial transcriptional signatures, and elevated normative expression of several genes, most prominently the immune checkpoint gene *LAG3*. These findings support a model in which brain metastasis susceptibility reflects interactions between circulating tumor cells and regional properties of the host tissue organized across multiple biological scales.

These findings extend longstanding theories regarding why certain regions of the brain are preferentially colonized by metastatic cells. The seed-and-soil hypothesis proposes that successful metastasis requires compatibility between disseminated tumor cells and the tissue environment they encounter.^11,12^ Mechanical theories instead emphasize the trapping of circulating tumor cells within microvasculature, particularly at vascular transition zones such as the gray-white matter junction.^13,55,56^ More recent studies have shown that seed-soil compatibility reflects complex interactions involving tumor cell surface markers,^57,58^ pre-metastatic niche formation,^59^ metabolic adaptation,^60–62^ blood-brain barrier remodeling,^63–65^ glial interactions,^66^ and immune evasion.^58,67^ Our analyses addressed the host-tissue side of three of these processes, metabolism, glial composition, and immune signaling, and each showed reproducible spatial associations with metastasis vulnerability. This suggests that at least some determinants of seed-soil compatibility vary regionally across the brain. The normal brain is increasingly recognized as exhibiting regional heterogeneity across multiple biological scales, including vascular,^68–70^ metabolic,^71^ cellular,^72^ transcriptomic,^36,39,73–75^ and immune^76,77^ processes. These intrinsic differences may collectively define region-specific niches that influence the permissiveness of the brain microenvironment to metastatic colonization.

Metastasis-prone regions showed greater perfusion, glucose metabolism, and mitochondrial density, and were supplied by smaller-caliber vessels. The perfusion and gray-white junction associations replicate prior work by Barrios et al.,^14^ and the additional vascular and metabolic associations suggest two complementary explanations. The first concerns delivery: regions with higher blood flow receive more circulating tumor cells, and smaller-caliber vessels favor their arrest, consistent with mechanical models of metastatic seeding.^13,55,56^ Because cerebral blood flow and glucose metabolism are tightly coupled,^78^ part of the metabolic association may reflect this shared vascular supply. The second concerns compatibility: brain metastatic cells adapt their metabolism to colonize the brain,^60–62^ and regions with high baseline glucose availability and oxidative capacity may better support these demands. Our data cannot distinguish these explanations, which are not mutually exclusive.

Prior studies of brain metastasis have focused primarily on the tumor microenvironment after colonization, showing that astrocytes,^79^ microglia,^80^ T cells,^81,82^ and other resident brain cells can support tumor growth, immune suppression, and treatment resistance.^47,66,83,84^ In contrast, we found that metastases preferentially localize to regions with low astrocyte and microglial transcriptional signatures in the healthy brain. These findings are not necessarily contradictory, because the two lines of evidence address different stages of disease. Before colonization, astrocytes and microglia can contribute to tissue surveillance and anti-tumor responses; after tumor entry, they may be co-opted into tumor-supportive states that promote growth and therapeutic resistance.^80,83^ Our normative maps capture the first of these stages: the baseline cellular landscape that circulating tumor cells encounter on arrival.

Experimental evidence supports a protective role for microglia at this stage. In a breast cancer model, mice genetically lacking microglia developed more brain metastases and showed impaired natural killer and T cell responses.^85^ Conversely, prophylactic microglial stimulation reduced the formation of new brain metastases.^86^ Human data are consistent with this pattern: the cerebellum, one of the most frequently colonized sites,^7–10^ has among the lowest microglial densities in both the healthy human^77,87^ and mouse brain.^88^ Together, these observations suggest that microglia, and possibly other resident glia, may restrain metastatic seeding in the healthy brain even as they support established tumors, which reconciles our findings with the pro-tumor roles reported after colonization.

Among the candidate genes, *LAG3* showed the strongest association of any canonical immune checkpoint, and its expression was also elevated in peritumoral tissue from surgically resected brain metastases.^47^ LAG-3 is an inhibitory immune checkpoint receptor that suppresses T-cell activation. It contributes to immunosuppressive tumor environments by inhibiting T-cell signaling and by modulating antigen-presenting cell function through MHC class II interactions.^51,89–91^ Brain metastases develop immunosuppressive niches characterized by impaired antigen presentation,^47^ T-cell exhaustion,^82,92^ and immune evasion,^67,84^ and *LAG3* expression has been reported within the brain metastasis microenvironment.^92–96^ Our findings extend this observation to the healthy brain: regions with high normative *LAG3* expression may provide an immunologically permissive “soil” for metastatic colonization before a tumor is established. LAG-3 may also contribute to the immunosuppressive environment that develops after colonization, through regulatory T-cell-mediated suppression and modulation of antigen-presenting cell maturation.^97,98^ LAG-3 is classically expressed by immune populations, including CD4+ and CD8+ T cells, natural killer cells, and dendritic cells;^99–103^ expression in neurons has also been reported^51,104^ but remains contested.^105^ Consistent with this, we detected *LAG3* in T cells, natural killer cells, and a subset of neurons in the Human Brain Cell Atlas,^39^ and expression in both the Allen Human Brain Atlas^36^ and the healthy pig brain^48^ was highest in the cerebellum, a region frequently colonized by metastases. Given the emergence of LAG-3 as a therapeutic target in melanoma^106^ and its ongoing investigation in glioblastoma,^107^ future studies should determine whether LAG-3 contributes causally to brain metastatic vulnerability and whether it represents a therapeutic opportunity in brain metastases.

Additional candidate genes provide complementary hypotheses. FGF9, whose spatial expression ranked among the most similar to that of brain metastasis vulnerability, is a fibroblast growth factor involved in cellular differentiation and tissue repair,^108^ and is known to be expressed in the brain,^109^ where it regulates development of the cerebellum.^110^ It has also been implicated in various cancer-related processes, including stimulating human brain tumor growth,^111,112^ and cancer cell metastasis.^113,114^ ALDH1A3, another top spatial correlate of BM vulnerability, is an aldehyde dehydrogenase implicated in tumor cell metabolism.^115^ Conversely, IL13RA2 was among the most negatively associated genes, meaning its normative expression could be interpreted as having a protective effect against BM colonization. Consistent with a protective role, experiments have shown that loss of IL13RA2 promotes brain metastasis.^116^ While our reported spatial associations do not establish causality, they nominate LAG3, FGF9, ALDH1A3, and IL13RA2, among others listed above, as candidate regulators of the pre-metastatic brain microenvironment for future mechanistic study.

Our findings also reveal that regional metastatic vulnerability is not entirely universal across tumor origins. Associations observed across lung, breast, and other carcinomas were broadly conserved, whereas melanoma diverged from this pattern. This divergence may reflect unique biological properties of melanoma metastasis, including greater heterogeneity of circulating tumor cells.^117,118^ Notably, melanoma is unique among the tumor types examined in arising from the neuroectoderm; this shared developmental lineage with the central nervous system may partly explain why melanoma metastasis spatial vulnerability diverges from carcinomas. Consistent with this possibility, melanoma-specific spatial associations were enriched for normative brain expression patterns related to axon guidance and synaptic signaling, suggesting that regional variation in neural developmental programs may contribute to the distinct spatial distribution of melanoma metastases.

Several aspects distinguish this work from prior studies of spatial brain metastasis patterns. This represents, to our knowledge, the largest analysis testing brain metastasis vulnerability against normative brain organization, and the first to incorporate independent discovery and validation cohorts to evaluate reproducibility of associations. Prior work by Barrios et al. established important relationships between metastasis location, gray-white matter junction proximity, and cerebral perfusion,^14^ but did not examine multimodal atlas-based associations spanning vascular, metabolic, cellular, and molecular organization. While Bao et al. explored relationships between metastasis distributions and normative brain features, they were limited by a smaller, single-institution cohort restricted to lung and breast cancer, lower-resolution gray matter parcellation approaches that excluded white matter and infratentorial regions including the cerebellum, and analytic methods designed for surface-based neuroimaging rather than volumetric lesion distributions.^119^ By incorporating the entire brain volume, including infratentorial structures, comparing to a more biologically comprehensive panel of normative atlases, and validating findings across large independent patient cohorts, our approach provides the most comprehensive assessment of the biological architecture correlating with metastatic susceptibility to date.

The present study also contributes to an emerging framework that uses multimodal, multiscale atlases of normative human brain organization to investigate CNS disease.^120–122^ Prior studies using the same normative gene expression dataset that we use herein have demonstrated that normative gene expression patterns recapitulate functional networks, structural connectivity, neurotransmitter distributions, cortical hierarchy, and regional vulnerability across neurological and psychiatric disorders.^74,123–139^ More recently, studies have begun linking normative gene expression patterns directly with lesion-related phenotypes, including neurodegeneration, movement disorders, and seizures.^37,140–144^ We build on this technique by using Allen Human Brain Atlas expression maps to construct cell-type-resolved atlases of the brain, which we then integrate directly with lesion locations. This allows us to identify associations between lesion locations at both the cellular level and at the level of individual gene expression. We show that, as with other lesion etiologies, the spatial distribution of brain metastases can be anchored to the molecular architecture of the healthy brain.

Several limitations warrant consideration. First, our analyses are inherently correlative and cannot establish causal relationships between regional brain properties and metastatic vulnerability. Second, normative atlases represent healthy brain organization and may not capture dynamic changes induced by aging, systemic cancer, prior therapies, or interactions with circulating tumor cells. Third, integration across imaging, vascular, and transcriptomic modalities is constrained by differences in spatial resolution, sampling, and biological scale. Because spatial autocorrelation, noise, and sampling density vary across modalities, correlation coefficients across biological domains should not be interpreted as directly comparable measures of effect magnitude. Fourth, transcriptomic and cell-type analyses rely on the Allen Human Brain Atlas, which is derived from six postmortem donors with variable sampling density and spatial coverage. Cell-type maps were further derived from canonical marker genes^39^ rather than direct measurements of cell abundance, so a lower regional signal may reflect either fewer cells or transcriptionally distinct populations that weakly express canonical markers; our approach cannot distinguish these possibilities. Fifth, the distribution of radiographically detectable metastases reflects the cumulative outcome of tumor cell arrest, extravasation, survival, and growth; therefore, our atlas cannot distinguish whether identified regional features influence initial seeding, successful colonization, or subsequent expansion, nor how these factors interact with tumor-specific determinants of metastatic behavior. Sixth, the Discovery cohort comprised patients treated with radiosurgery, which may underrepresent large or surgically resected metastases. Segmentation methods also differed across cohorts (radiosurgery contours, manual expert segmentation, and automated segmentation with manual correction); replication despite these differences supports the generalizability of the associations, although method-specific bias cannot be fully excluded. Finally, spatial normalization of brains containing mass lesions is inherently imperfect, and despite modern deformable registration methods, residual anatomical distortion may introduce uncertainty into voxel-level localization estimates.

## CONCLUSIONS

Brain metastases preferentially localize to regions of the healthy brain with distinct vascular, metabolic, cellular, and molecular features, and these associations replicated within a large independent cohort. Metastasis-prone regions were highly perfused and metabolically active, showed low astrocyte and microglial transcriptional signatures, and showed high normative expression of the immune checkpoint gene *LAG3*. These patterns were shared across carcinoma subtypes but diverged in melanoma. Together, these findings suggest that regional vulnerability to brain metastasis is shaped in part by the pre-existing organization of the brain itself, the “soil” that circulating tumor cells encounter on arrival. If confirmed experimentally, this framework could inform region-specific surveillance and nominate the brain’s regional immune landscape, including LAG-3, as a target for preventing or treating brain metastases.

## Data Availability

Normative atlas sources are publicly available, detailed in Supplementary Table S2. The NYUMets and UCSF datasets are publicly available. BWH/DFCI and Zurich data are available from the corresponding authors upon reasonable request.

https://nyumets.org/

https://imagingdatasets.ucsf.edu/dataset/1

## Ethics

The NYUMets and UCSF datasets are publicly available and de-identified;^17,22^ their analysis involved no patient contact, consent, or identifiable data and did not require institutional review board (IRB) approval. Retrospective use of the BWH/DFCI and University of Zurich data was approved by the respective institutional review boards or ethics committees, with a waiver of informed consent, and was conducted in accordance with the Declaration of Helsinki.

## Data Availability

Normative atlas sources are publicly available, detailed in Supplementary Table S2. The NYUMets and UCSF datasets are publicly available.^15,17,145^ BWH and Zurich data are available from the corresponding authors upon reasonable request.

## Authorship

J.I.T. conceived the study, performed the analyses, and wrote the manuscript. A.A., A.Y.F., and K.B. contributed to data collection and analysis. A.A.A., N.L., H.E., and R.M. assembled and curated the BWH/DFCI cohort. J.P. performed automated segmentation of the BWH/DFCI lesions under the supervision of R.Y.H. A.Y.S. segmented the Zurich cohort lesions. F.W., M.G., and M.W. provided the Zurich cohort. M.E.P., E.K.O., F.L.W.V.J.S., and D.K. supervised the work, contributed to interpretation, and provided data. All authors reviewed and approved the final manuscript.

## SUPPLEMENTARY METHODS

### Cohort Details and Lesion Segmentation

In the Discovery cohort (n = 1,406), patients were drawn from the NYUMets dataset.^17,146^ Primary tumor etiologies included lung, breast, melanoma, genitourinary, and gastrointestinal malignancies; detailed cohort characteristics and data generation procedures have been described previously.^17^ Metastasis segmentations were the tumor target contours delineated for radiosurgery planning, extracted from the corresponding DICOM RTSTRUCT files. Baseline T1-weighted MRI scans and segmentations were normalized to the MNI152 template distributed with FSL using EasyReg, a deep learning-based deformable registration tool implemented within FreeSurfer.^18,147,148^

In the Validation cohort (n = 1,209), the Brigham and Women’s Hospital (BWH) dataset included 702 patients drawn from a cohort of 1,453 patients with brain metastases.^19^ Clinical data, including primary tumor type, were obtained from that study, and pretreatment MRI scans were retrieved for [inclusion criteria; CONFIRM]. Metastases were segmented using an automated deep learning algorithm [SPIRS; CITE Patel et al. 2023, CONFIRM], and each segmentation was manually reviewed and corrected as needed. The University of Zurich dataset included 193 patients, with manual lesion segmentations and primary tumor data obtained from a prior study.^21^ The UCSF dataset included 314 patients with expert segmentations from a publicly available Gamma Knife radiosurgery dataset (https://imagingdatasets.ucsf.edu/dataset/1), including patients treated for lung, breast, melanoma, genitourinary, and gastrointestinal malignancies.^22,145^

### Normative Atlases of Brain Physiology, Gene Expression, and Cell Types

We assembled normative volumetric atlases spanning microstructural organization, vascular architecture, and metabolism (Supplementary Table S2). Microstructural atlases included synaptic density,^23^ myelin,^24^ and gray and white matter probability.^25,26^ To estimate gray-white junction probability, we generated a composite atlas by multiplying voxelwise gray and white matter probability values. Vascular atlases included cerebral blood flow,^27^ cerebral blood volume,^28^ medium-to-large arterial structure density,^29^ vessel radius,^30,31^ venous density, vessel diameter, and vessel torsion.^32,33^ Metabolic atlases included mitochondrial density^34^ and glucose metabolism.^35^ All atlases were distributed in MNI152 space and resampled to 2 mm isotropic resolution using default continuous interpolation in Nilearn.^149^

Transcriptomic maps for 15,633 genes were derived from the Allen Human Brain Atlas (AHBA),^36^ a normative dataset built from postmortem microarray profiling of six donor brains across 3,702 sampled regions. We used voxelwise expression maps from prior work,^37^ (N. Nolan, unpublished manuscript) generated using the abagen toolbox^38^ with standard recommended parameters:^150^ 0.5 intensity-based filtering, bidirectional hemispheric mirroring, differential stability probe selection, nearest-neighbor interpolation with donor-wise averaging, and cross-donor robust sigmoid normalization, followed by voxelwise z-scoring.

For each of the 31 cell-type superclusters defined by Siletti et al.,^39^ we retained the marker genes reported in that publication’s Supplementary Table S3 that were represented in the AHBA (median 23 genes per cell type), retrieved the corresponding AHBA expression maps, and computed the first principal component across each cell type’s marker genes. Principal component analysis was used instead of voxelwise averaging because normalized expression values are comparable within a gene across voxels but not across genes; the first principal component instead captures spatial structure shared across the marker set.

### Simulation of Null Lesion Distributions and Spatial Permutation Testing

To address the concern that if metastases occurred randomly throughout the brain, atlas correlations may arise simply because lesions have finite size and must lie within brain anatomy, we used permutations to test this null hypothesis (Fig. S10). To generate empirical null distributions for spatial permutation testing, we simulated metastasis distributions directly within the brain volume, as previously described^15^ (medRxiv, doi: 10.64898/2026.06.29.26356888).

This approach was designed to approximate random spatial relocation of lesions while (1) avoiding invalid lesion placements outside brain boundaries, and (2) avoiding the central-brain bias that would otherwise be introduced by excluding such invalid placements. Because volumetric lesion maps cannot be permuted using surface-based approaches such as spherical rotation (“spin tests”), which are valid only for cortical surface data, this voxelwise simulation strategy was used in their place.

#### Single-lesion growth procedure

Each simulated lesion was generated by first selecting a seed voxel from within the brain mask (excluding ventricles), with all valid voxels treated as equally likely to be selected. Beginning at this seed, the lesion was grown one voxel at a time by iteratively incorporating a neighboring, not-yet-occupied voxel, with selection weighted toward voxels proximal to the lesion origin. This produced compact, approximately convex lesions consistent in morphology with observed metastases. These simulated lesions exhibited similar sphericity (0.76 ± 0.11 vs. 0.75 ± 0.09) and convexity (1.58 ± 0.68 vs. 1.39 ± 0.55) as the observed NYUMets lesions. Growth continued until the lesion reached a predefined target volume, drawn from the empirical volume distribution of the observed cohort. This procedure was repeated independently to generate as many lesions as desired for a given simulated map.

#### Generation of the null ensemble and multiple comparison correction

Using this procedure, we generated 5,000 permuted lesion maps, each matched exactly to the observed cohort in both the number of simulated lesions (e.g., n = 6,011 in the Discovery cohort) and the volume of each individual lesion. Within each permutation, all voxels within the brain mask were treated as equally probable seed and growth locations (i.e., no voxelwise prior was applied), reflecting the null hypothesis of spatially unbiased metastatic distribution. For each simulated map, all lesions were superimposed to generate a simulated frequency map, exactly as done for the observed data. Statistics of interest (e.g., spatial correlations with normative atlases) were recalculated for each of the 5,000 simulated maps to generate an empirical null distribution, against which the true observed statistic was compared. Two-sided empirical P values were calculated as the proportion of simulated statistics with a correlation magnitude equal to or exceeding the observed value and were corrected for multiple comparisons within each a priori biological category (microstructural, vascular, metabolic, cellular, and transcriptomic) using the Benjamini-Hochberg false discovery rate (FDR) procedure^151^ (α = 0.05). Atlases significant in the Discovery cohort were tested in the Validation cohort using the same permutation procedure, with FDR correction applied across the atlases significant in the Discovery cohort for that category.

#### Validation against the observed metastasis distribution

As a positive control, we additionally generated simulated lesion maps in which voxel selection, both for lesion seeding and lesion growth, was weighted by the observed empirical metastasis frequency map, rather than treated as spatially uniform. This analysis served as an implementation check demonstrating that the lesion-generation procedure itself does not distort an imposed spatial prior. The resulting realistic simulated frequency map closely resembled the true observed metastasis frequency map (spatial correlation r = 0.80), confirming that the simulation framework faithfully captures large-scale patterns of metastatic spatial distribution when supplied with an appropriate prior.

### Candidate Gene Validation, Disease Specificity, Covariate Adjustment, and GSEA

In the two-part candidate gene selection criteria, a gene had to rank among the top 10 or bottom 10 spatial correlations with the Discovery metastasis map and exceed a two-tailed significance threshold in the Validation cohort (α = 0.025, corresponding to the 98.75th and 1.25th percentiles of the validation null distribution). Nominated candidates were then confirmed against the spatial permutation framework described above (FDR p < 0.001).

To assess disease specificity, we compared metastasis locations against lesion masks from publicly available atlases of stroke,^40^ meningioma,^41^ focal cortical dysplasia,^42^ and multiple sclerosis,^43,44^ alongside randomly generated lesions matched by the permutation procedure used to construct null metastasis maps. For each candidate gene, normative expression values were averaged within each patient’s lesion mask, and expression at metastasis locations was compared with expression at other lesion locations using the Mann-Whitney U test, with FDR correction across candidates (all FDR p < 0.001). This analysis reflects normative regional gene expression at these locations, not expression within the lesions themselves.

To confirm that candidate associations were not driven by the physiological features identified in Figure 2, we fit an ordinary least squares model (statsmodels^152^) predicting voxelwise Discovery metastasis frequency from the reproducible microstructural, vascular, and metabolic covariates, and re-correlated all gene expression maps against the model residuals. All candidate genes remained highly ranked (top or bottom 25) in this covariate-adjusted analysis.

Gene set enrichment analysis (GSEA)^45^ was performed on the full ranked list of gene-map correlations from the Discovery cohort using the Gene Ontology Biological Process database^153,154^ and cell-type marker sets,^39^ with FDR correction (_α_ = 0.05). Gene sets significant for positive or negative enrichment were re-evaluated using the corresponding ranked list from the Validation cohort.

### Supratentorial Sensitivity and Etiology-Stratified Analyses

To test whether candidate associations depended on differences between the cerebellum and cerebrum, we repeated the spatial correlation analysis within a supratentorial mask based on the Harvard-Oxford atlas.^155,156^ The Discovery and Validation metastasis frequency maps, the 17 candidate gene expression maps, and the five reproducible cell-type maps were restricted to voxels within this mask, and Pearson correlations were recomputed. Significance was assessed with the spatial permutation framework using 5,000 null metastasis maps restricted to the same mask, with FDR correction applied separately within the gene and cell-type categories.

For etiology-stratified analyses, all reproducible physiological, cell-type, and gene-level associations were recalculated using the etiology-stratified atlases for lung, breast, other carcinomas, and melanoma. For melanoma, we additionally applied the candidate gene criteria and GSEA^45^ to the melanoma-specific Discovery and Validation maps. Leading-edge genes were defined as those shared across multiple reproducibly enriched gene sets and ranked in the top decile of melanoma-associated genes in both cohorts.

### Immune Checkpoint and *LAG3* Validation

For the six canonical immune checkpoint genes (*PDCD1* [PD-1], *CD274* [PD-L1], *CTLA4*, *LAG3*, *HAVCR2* [TIM-3], and *TIGIT*),^46^ AHBA expression maps were spatially compared with the Discovery and Validation metastasis maps; a combined map pooling both cohorts was also used to generate a single summary statistic. We assessed *LAG3* expression in an open spatial RNA-sequencing dataset of surgically resected non-small cell lung cancer brain metastases,^47^ comparing peritumoral microenvironment samples (n = 20) with control tissue samples (n = 7) using Welch’s two-sample t-test (α = 0.05). *LAG3* expression was higher in peritumoral than control tissue (Welch’s *t* = 3.28, df = 19.51, *p* = 0.0038). We additionally quantified *LAG3* normalized transcripts per million across pig brain regions from an open dataset.^48^ Finally, we characterized the cellular distribution of *LAG3* in the Human Brain Cell Atlas^39^ (approximately 3.3 million neuronal and non-neuronal cells) as the percentage of cells within each cell type expressing *LAG3*. All statistical analyses were performed in Python (pandas, numpy, nilearn, scipy, statsmodels, nibabel, abagen) and R (fgsea).

