## Supplementary Figures for "Regional Vulnerability to Brain Metastases Reflects Structural, Metabolic and Genetic Organization of the Healthy Brain"

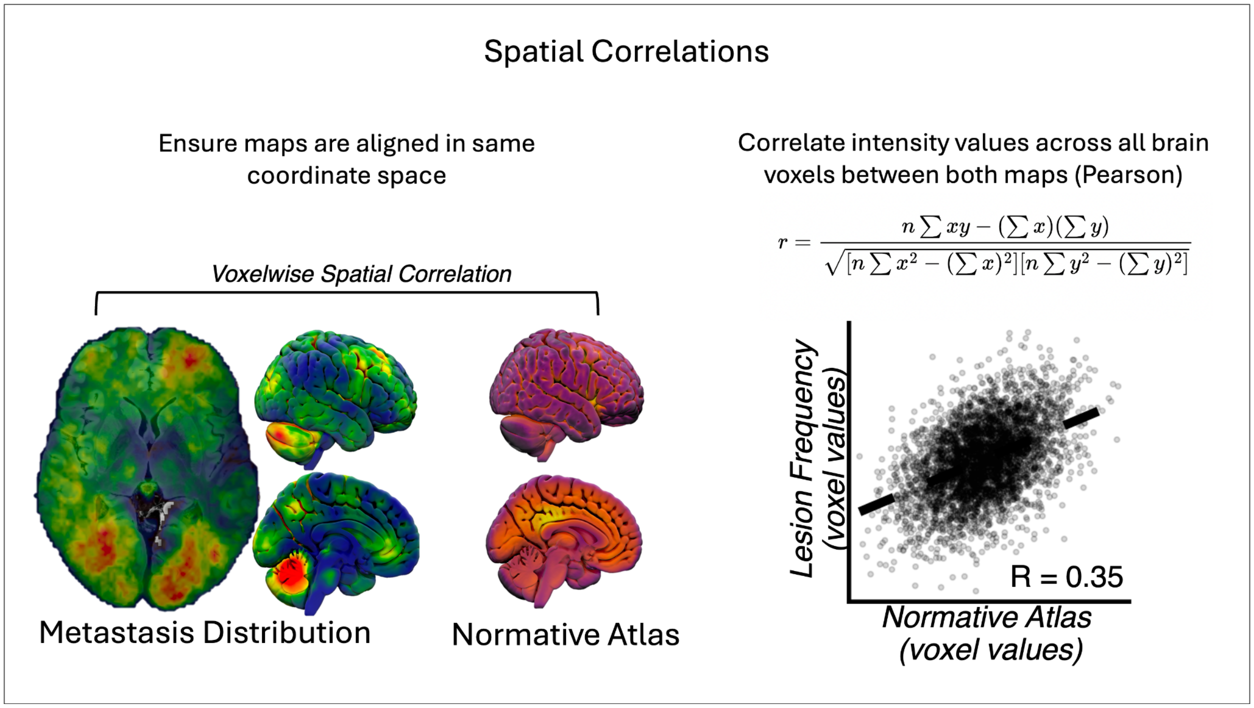


**Figure S1. Voxelwise spatial correlation methodology.** Metastasis distribution maps and normative brain atlases were first spatially normalized to a common coordinate space (MNI152) to ensure voxelwise correspondence. Intensity values were extracted at each brain voxel from both maps and correlated using Pearson's r, yielding a single correlation coefficient summarizing the spatial relationship between metastasis distribution and the normative atlas across the entire brain. This procedure was applied independently to each normative atlas and repeated separately in the Discovery and Validation cohorts.

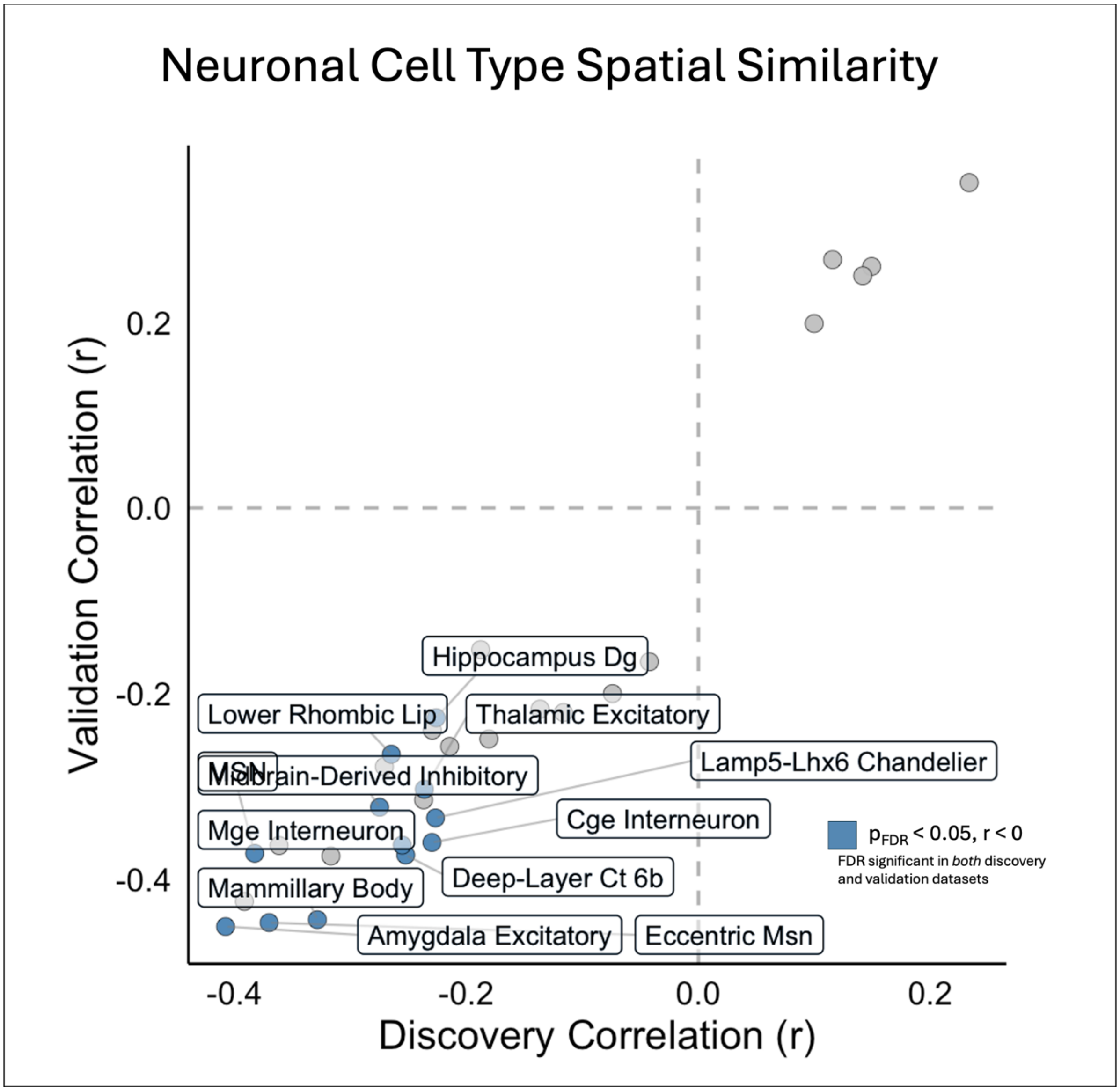


**Figure S2. Spatial correlation between metastasis distribution and neuronal cell-type atlases.** Spatial correlation between metastasis distribution and cell-type atlases for all neuronal cell-type superclusters from the Human Brain Cell Atlas, shown for the Discovery (x-axis) and Validation (y-axis) cohorts. Blue points indicate neuronal cell types with a statistically significant (FDR-corrected $P < 0.05$), reproducible inverse spatial correlation with metastasis distribution in both cohorts; gray points indicate cell types not meeting this threshold. Labeled points correspond to the neuronal superclusters exhibiting the strongest reproducible inverse associations, including mammillary body neurons, medium spiny neurons (MSN), eccentric MSN, amygdala excitatory neurons, midbrain-derived inhibitory neurons, and lower rhombic lip-derived neurons.

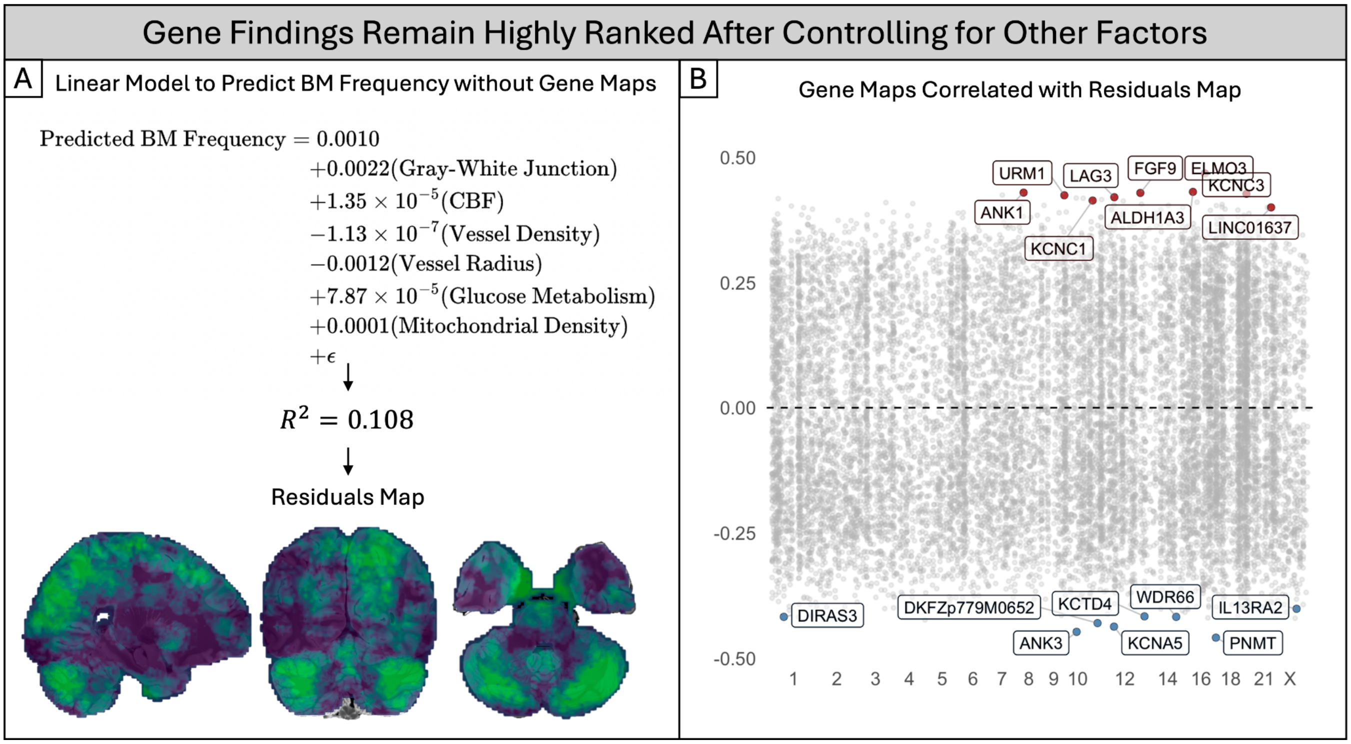


**Figure S3. Candidate gene associations with metastasis vulnerability remain robust after accounting for microstructural, vascular, and metabolic covariates.**

**(A)** A linear model was fit to predict voxelwise metastasis distribution using the microstructural, vascular, and metabolic normative atlases identified in Figure 2 (gray-white junction, cerebral blood flow, vessel density, vessel radius, glucose metabolism, and mitochondrial density), without including any gene expression maps. The fitted model explained a modest proportion of variance in metastasis distribution ($R^2 = 0.108$), and the residuals from this model (representing variation in metastasis distribution not explained by these physiological covariates) were retained as a residual map (bottom, axial, coronal, and sagittal views shown).

**(B)** Spatial correlation between the residual map from (A) and all 15,633 gene expression maps from the Allen Human Brain Atlas, organized by chromosome. Genes are plotted analogously to Figure 5A, but correlated against the covariate-adjusted residual map rather than the raw metastasis distribution map. Labeled genes correspond to the same top- and bottom-ranked candidates identified in the primary analysis (Figure 5), including LAG3, ALDH1A3, and FGF9, indicating that these associations are not simply secondary to the physiological niche characterized in Figure 2 and persist after statistically accounting for it.

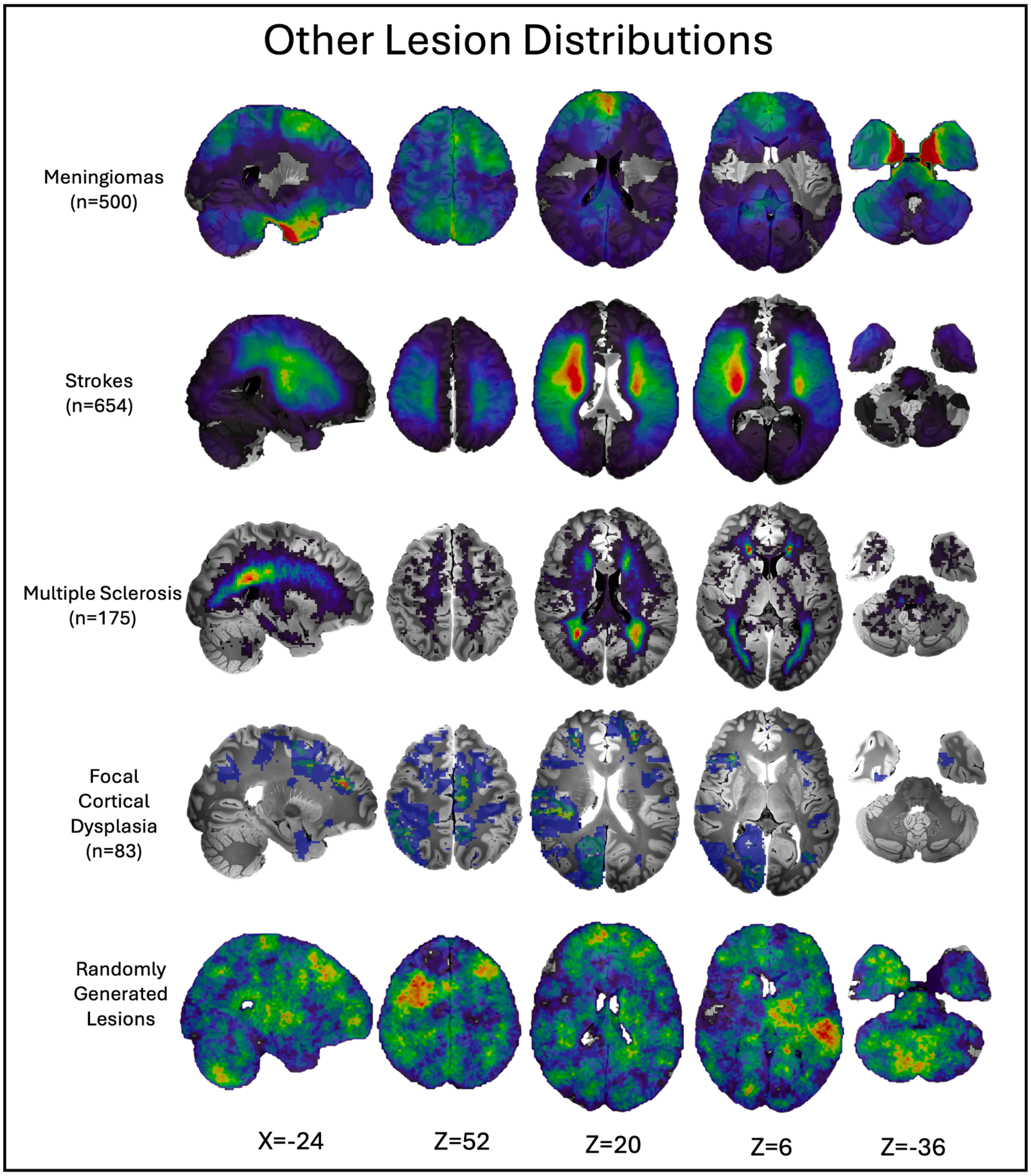


**Figure S4. Voxelwise lesion frequency maps for other pathologies.** Representative sagittal (X = -24), axial (Z = 52, 20, 6), and cerebellar (Z = -36) sections showing voxelwise lesion frequency distributions for meningioma (n = 500), stroke (n = 654), multiple sclerosis (n = 175), focal cortical dysplasia (n = 83), and randomly generated lesions. These distributions correspond to the other lesion types referenced in Figure 5C.

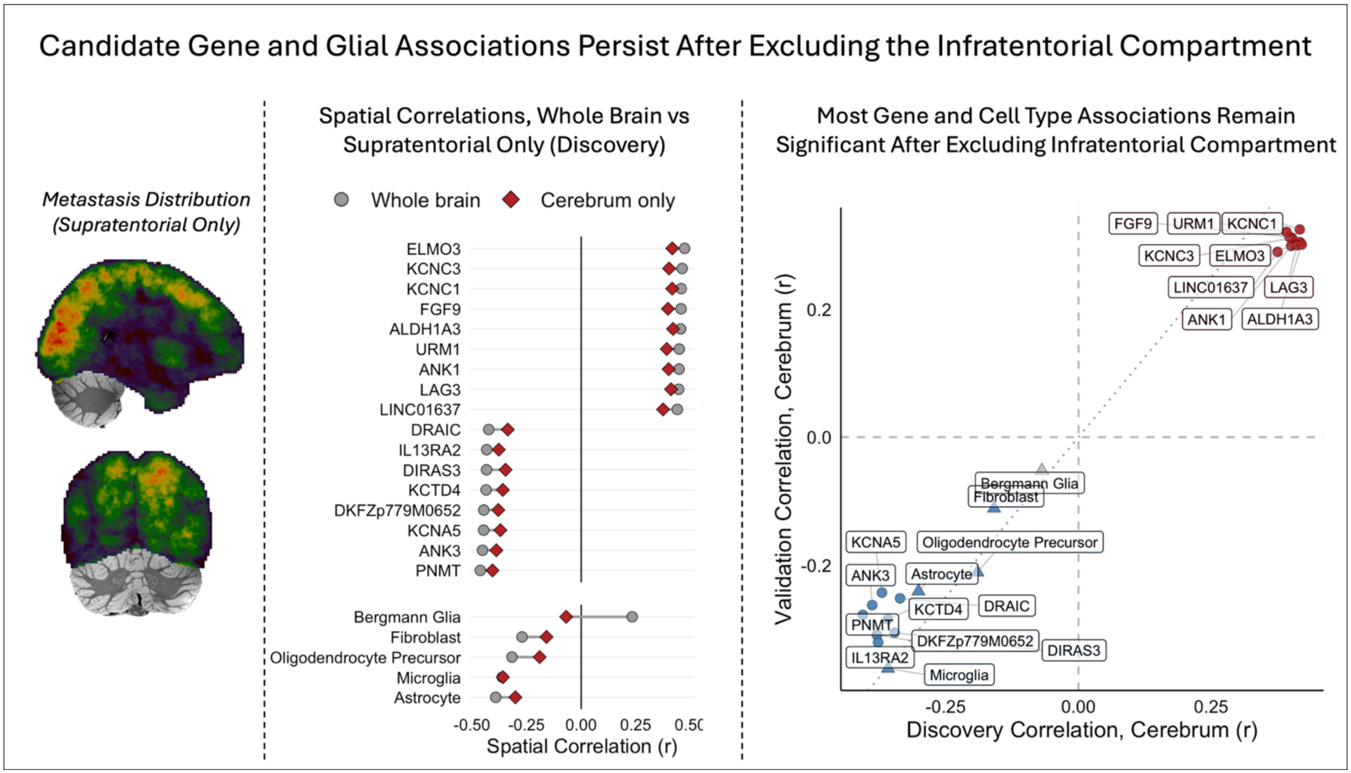


**Supplementary Figure S5. Candidate gene and cell-type associations after excluding the infratentorial compartment.** (A) Discovery metastasis frequency map restricted to the supratentorial compartment, shown in sagittal and coronal views. (B) Spatial correlations between the Discovery metastasis frequency map and each candidate gene expression map and reproducible cell-type map, computed across all brain voxels (gray circles) or supratentorial voxels only (red diamonds). (C) Supratentorial-only spatial correlations in the Discovery and Validation cohorts. Red and blue indicate positive and negative associations that were significant in both cohorts (FDR-corrected $P < 0.05$, spatial permutation test, 5,000 permutations); gray indicates associations that did not replicate. Circles, genes; triangles, cell types.

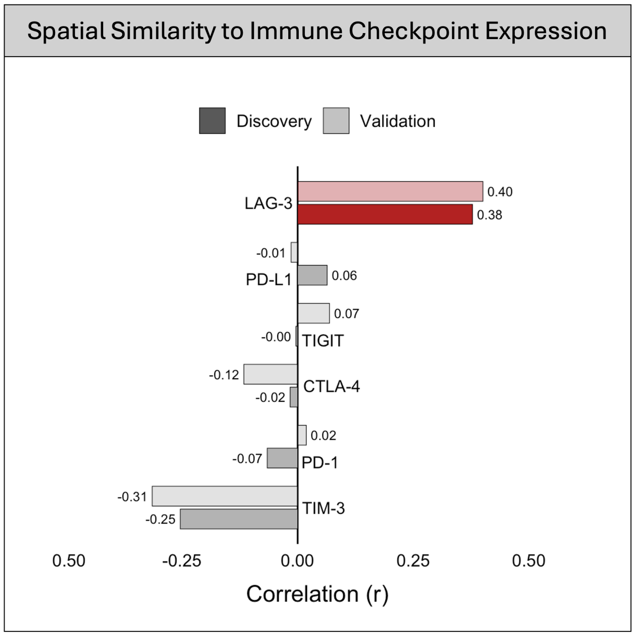


**Figure S6. Spatial Correlations with Immune Checkpoint Genes.** Spatial correlations between discovery and validation metastasis distribution maps with six immune checkpoint genes (PD-1, PD-L1, LAG-3, CTLA-4, TIM-3, TIGIT). Only LAG3 shows consistent positive associations with metastasis distribution.

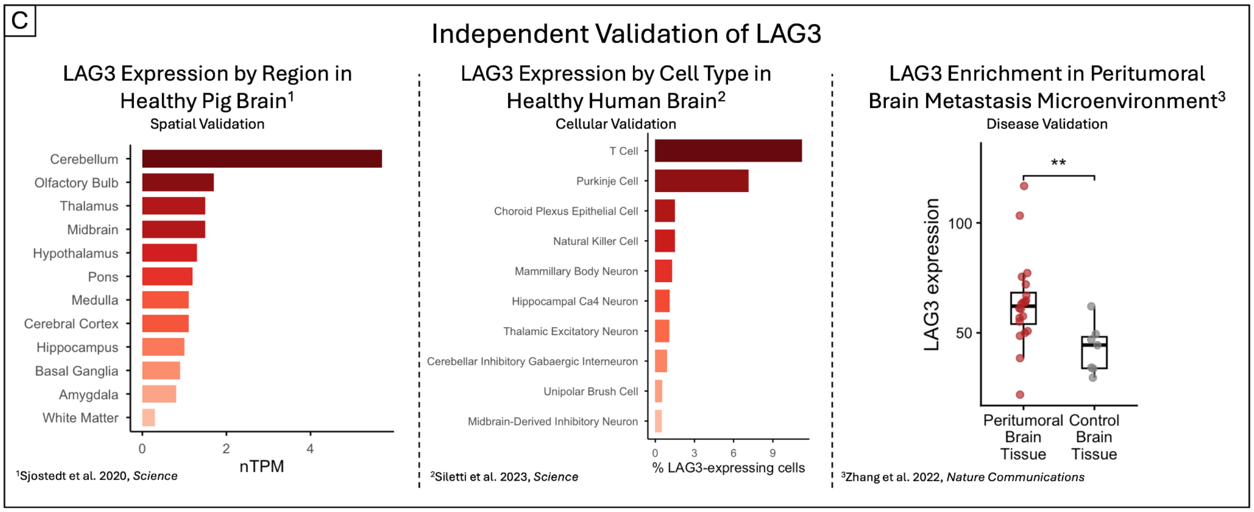


**Figure S7. Independent validation of LAG3 across species, cell type, and disease state.** Left, LAG3 expression across regions of the healthy pig brain, quantified in normalized transcripts per million (nTPM).[1] Middle, LAG3 expression across cell types in the healthy human brain, quantified as the percentage of LAG3-expressing cells per cell type.[2] Right, LAG3 expression in peritumoral brain metastasis tissue compared to control brain tissue (**$p < 0.01$).[3]

[1]Sjöstedt et al., 2020, *Science* [2]Siletti et al., 2023, *Science*. [3]Zhang et al., 2022, *Nature Communications*.

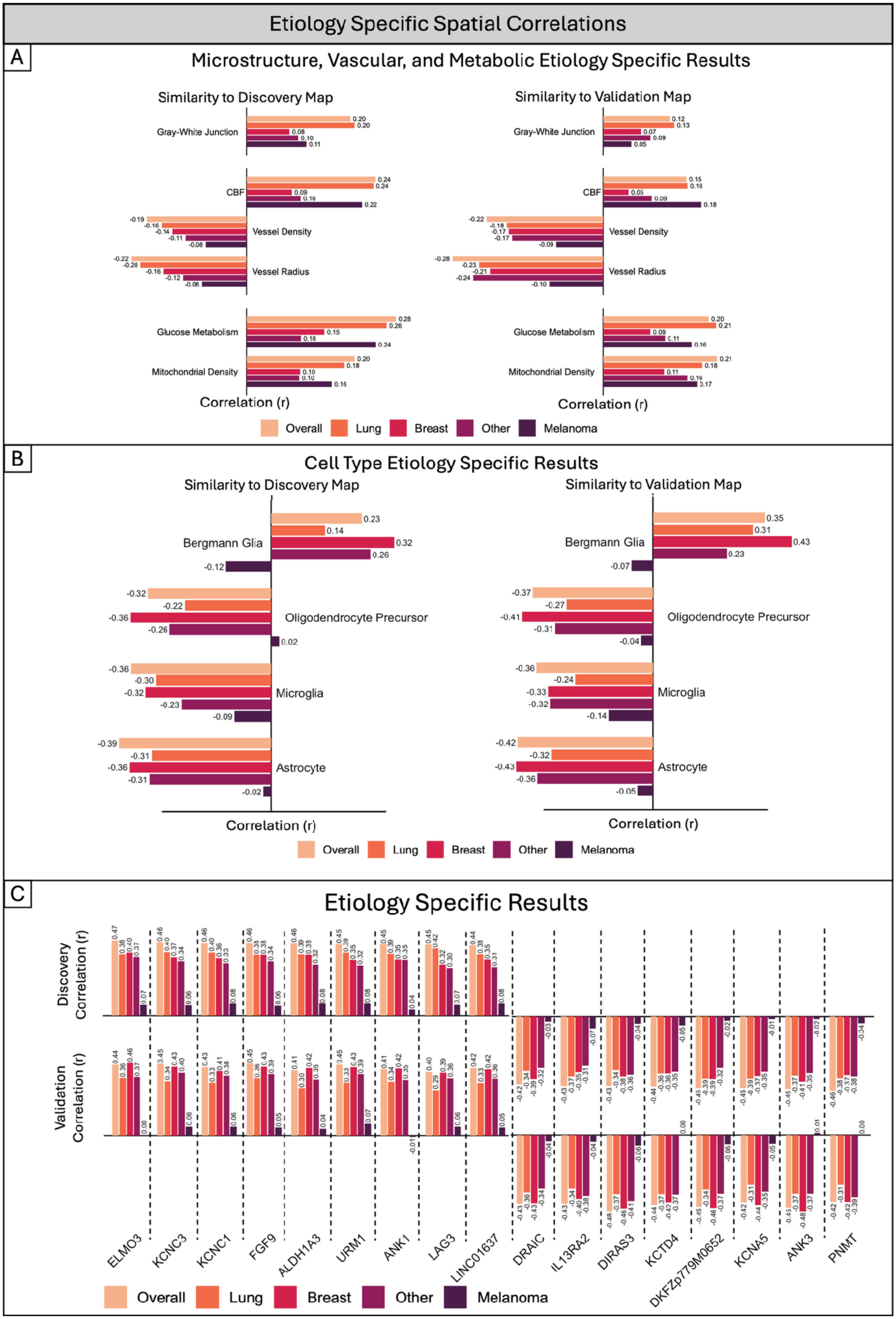

Etiology Specific Spatial Correlations
A
Microstructure, Vascular, and Metabolic Etiology Specific Results
Similarity to Discovery Map
Similarity to Validation Map
Gray-White Junction
CBF
Vessel Density
Vessel Radius
Glucose Metabolism
Mitochondrial Density
Correlation (r)
Overall
Lung
Breast
Other
Melanoma
B
Cell Type Etiology Specific Results
Similarity to Discovery Map
Similarity to Validation Map
Bergmann Glia
Oligodendrocyte Precursor
Microglia
Astrocyte
Correlation (r)
Overall
Lung
Breast
Other
Melanoma
C
Etiology Specific Results
Discovery Correlation (r)
Validation Correlation (r)
ELMO3
KCNC3
KCNC1
FGF9
ALDH1A3
URM1
ANK1
LAG3
LINC01637
DRAIC
IL13RA2
DIRAS3
KCTD4
DKFZp779M0652
KCNA5
ANK3
PNMT
Overall
Lung
Breast
Other
Melanoma

**Figure S8. Etiology-specific spatial correlations recapitulate the overall metastasis vulnerability niche, with melanoma as a consistent outlier.** For each association identified as significant and reproducible in the pooled (all-etiology) analysis, spatial correlations were recalculated separately within etiology-stratified metastasis distribution maps (lung, breast, other, and melanoma), alongside the original overall correlation for comparison.

**(A)** Microstructural, vascular, and metabolic atlas correlations, shown for the Discovery (left) and Validation (right) maps. Direction and magnitude of association were broadly consistent across all etiologies, including melanoma.

**(B)** Cell-type atlas correlations, shown for the Discovery (left) and Validation (right) maps. Lung, breast, and other carcinomas showed comparable direction and magnitude of association across all cell types; melanoma-specific correlations were consistently attenuated or reversed.

**(C)** Correlations for individual candidate genes, shown as gene expression correlation (top) and spatial correlation with metastasis distribution (bottom) for each etiology. As in (B), lung, breast, and other carcinomas tracked closely with the overall pooled correlation for each gene, while melanoma showed consistently weaker or divergent associations across nearly all candidates, including LAG3.

Together, these etiology-stratified analyses indicate that the physiological and cell-type associations underlying the metastasis vulnerability niche are largely shared across the carcinoma subtypes examined, whereas melanoma metastases diverge.

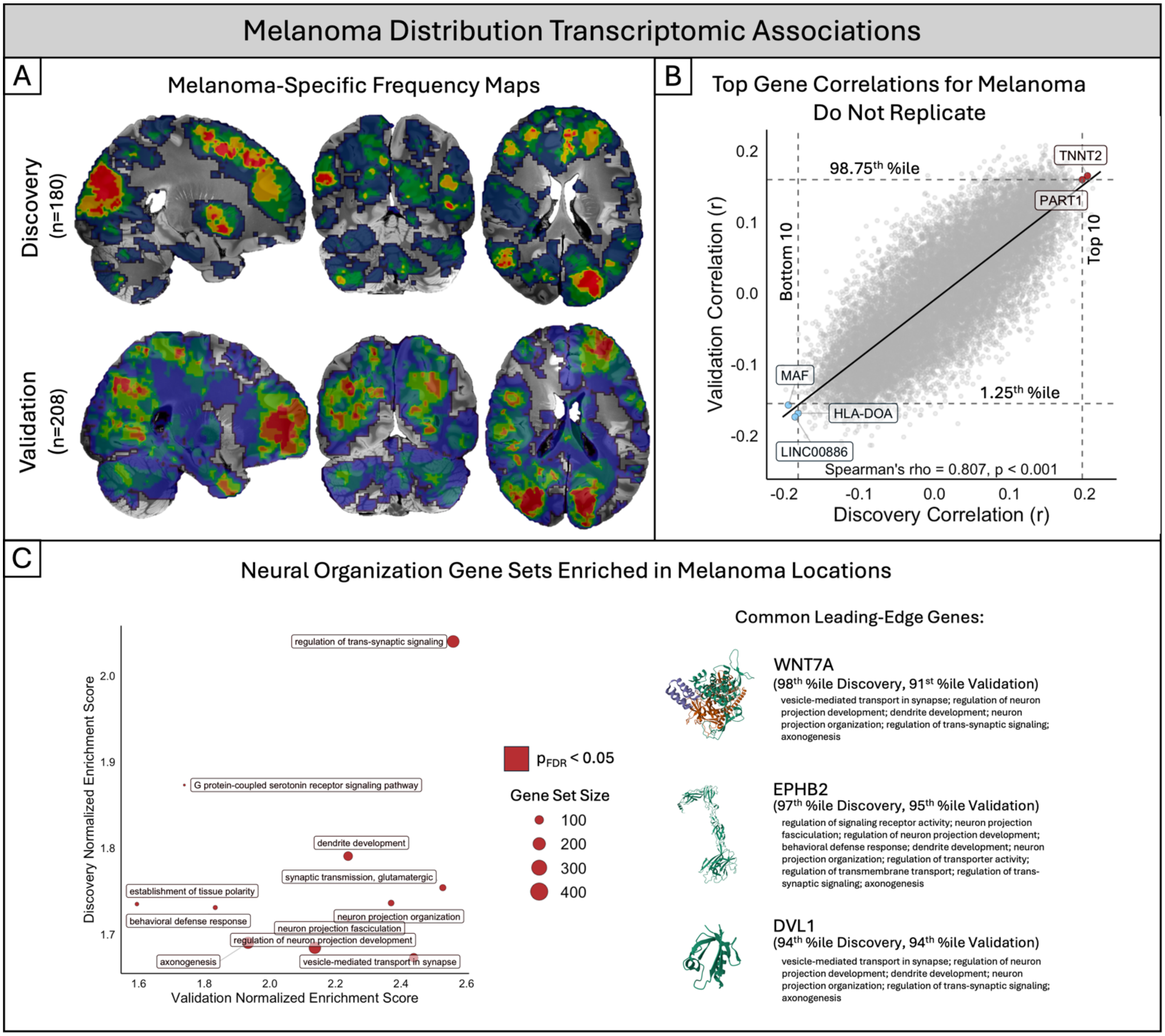


**Figure S9. Melanoma-specific spatial correlations implicate developmental and axon-guidance signaling despite lack of individual gene replication.**

**(A)** Melanoma-specific metastasis distribution maps in the Discovery (n = 133) and Validation (n = 208) cohorts, shown in sagittal, coronal, and axial views.

**(B)** Spatial correlation between melanoma-specific metastasis distribution and gene expression across all Allen Human Brain Atlas genes, shown for the Discovery (x-axis) and Validation (y-axis) cohorts. Although overall correlation strength between cohorts was substantial (Spearman's rho = 0.620, $p < 0.001$), individual top- and bottom-10 candidate genes identified in Discovery did not consistently replicate in Validation using the same top/bottom-10 and percentile thresholds applied to the pooled analysis (Figure 5A); only one gene (LOC101927502) met criteria in both cohorts.

**(C)** Gene set enrichment analysis (GSEA) of the ranked gene-metastasis spatial correlations. Gene sets related to neural organization and synaptic signaling—including regulation of trans-synaptic signaling, dendrite development, and axonogenesis—were reproducibly enriched across Discovery and Validation cohorts (point size indicates gene set size; red indicates FDR-significant reproducibility in both cohorts). To identify candidate genes underlying these convergent pathways, we examined leading-edge genes shared across multiple enriched ontologies that also ranked within the top decile of melanoma-associated genes in both cohorts. Three genes met these criteria: WNT7A (r = 0.088, 98th percentile Discovery; r = 0.120, 91st percentile Validation), EPHB2 (r = 0.080, 97th percentile Discovery; r = 0.133, 95th percentile Validation), and DVL1 (r = 0.074, 94th percentile Discovery; r = 0.132, 94th percentile Validation).

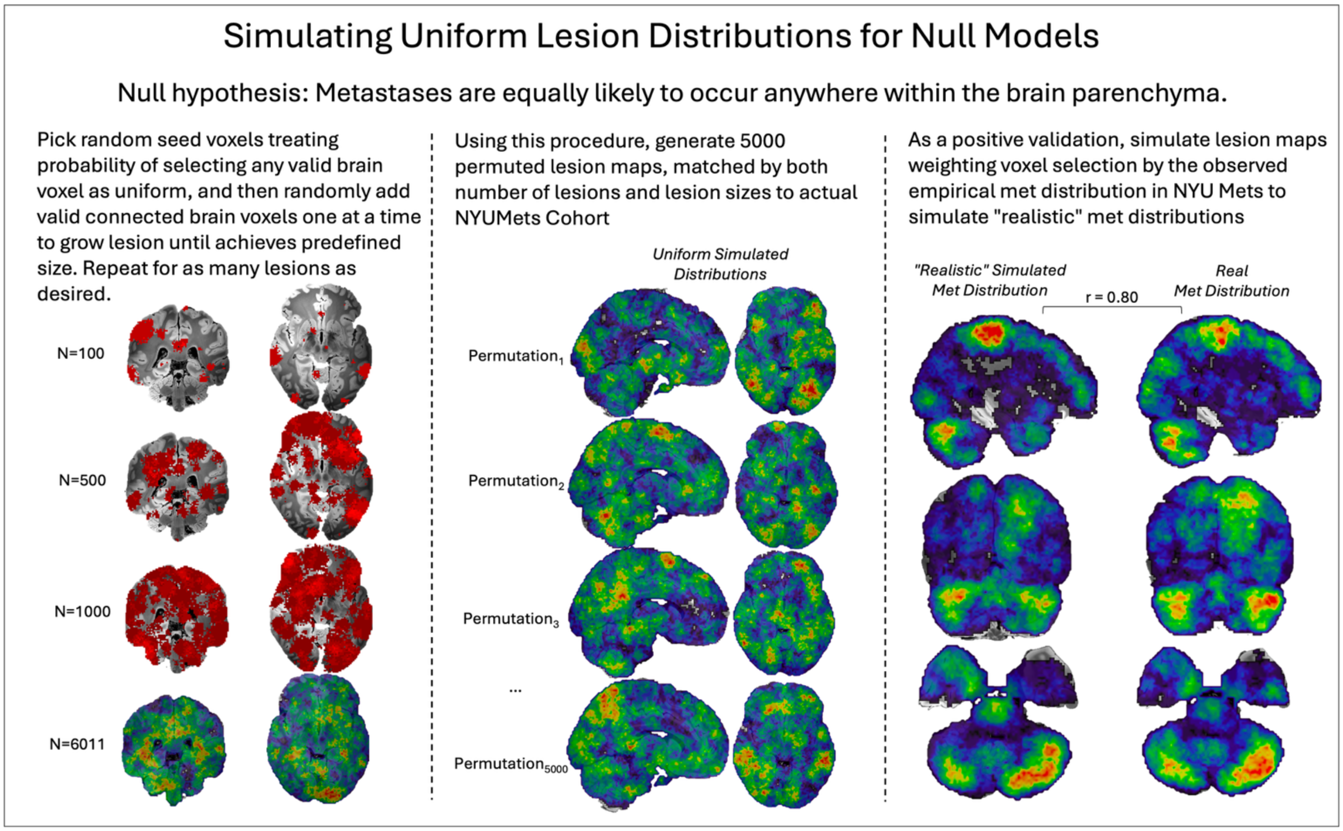


**Figure S10. Spatial permutation null model.**
Under the null hypothesis, metastases are equally likely to arise anywhere in the brain parenchyma. Each simulated lesion starts at a uniformly random parenchymal seed voxel and grows by adding randomly chosen adjacent voxels until it reaches a set size. Example maps show 100 to 6,011 simulated lesions. We generated 5,000 permuted lesion maps, each matched to the NYUMets cohort in lesion number and size, to serve as null distributions. As a positive control, weighting seed selection by the observed NYUMets distribution produced a map that closely matched the real metastasis frequency map ($r$ = 0.80).
