## Supplementary Tables for "Regional Vulnerability to Brain Metastases Reflects Structural, Metabolic and Genetic Organization of the Healthy Brain"

#### Supplementary Table S1.

#### Characteristics of the brain metastasis datasets used to generate metastasis distribution maps.

| | Discovery | Validation | | | | All datasets |
|---|---|---|---|---|---|---|
| **Characteristic** | **NYU** | **UCSF** | **BWH** | **Zurich** | **Combined** | **Combined** |
| N patients | 1406 | 314 | 702 | 193 | 1209 | 2615 |
| Female, % | NR | 59.6 | 59 | 49 | 57.6 | NR |
| Age at diagnosis, y, mean (SD) | 61.6 (12.8) | 61.4 (11.7) | 60 (12) | 60 (12) | 60.4 (11.9) | 61.0 (12.4) |
| **Primary tumor, %** | | | | | | |
| Lung | 53 | 40 | 42 | 41 | 41 | 48 |
| Melanoma | 13 | 14 | 18 | 19 | 17 | 15 |
| Breast | 14 | 23 | 18 | 11 | 18 | 16 |
| Other | 20 | 23 | 23 | 29 | 24 | 21 |
| **Lesion burden** | | | | | | |
| Total metastases, n | 6011 | 2629 | 2494 | 344 | 5467 | 11478 |
| Metastases per patient, mean (SD) | 4.28 (4.90) | 8.37 (12.92) | 3.6 (5.4) | 1.9 (1.6) | 4.57 (8.13) | 4.41 (6.59) |
| Metastasis volume, $cm^3$, mean (SD) | 1.58 (4.38) | 0.81 (2.86) | 2.9 (8.6) | 13.4 (23.0) | 2.55 (8.93) | 2.05 (6.95) |

NR, not reported; BWH, Brigham and Women's Hospital; SD, standard deviation.
Combined values are pooled from cohort-level means and SDs; metastasis volume is pooled across lesions. Female % for all datasets is not reported because sex is unavailable for NYU.

### Supplementary Table S2. Normative brain atlases used in spatial correlation analyses.

#### MICROSTRUCTURE ATLASES *(5 maps)*

| Atlas | Description | Source / DOI |
|---|---|---|
| Synaptic density | Normative map of synaptic density derived from PET imaging. | 10.1177/0271678X17724947 |
| Myelin (T1w/T2w Ratio) | Normative map of myelin content. | 10.1038/nature18933 |
| Gray-white matter interface probability | Derived map generated by voxelwise multiplication of gray matter and white matter probability maps (below), yielding an index of gray-white junction likelihood at each voxel. | 10.1016/S1053-8119(09)70884-5<br>10.1016/j.neuroimage.2010.07.033 |
| Gray matter probability | Voxelwise probability map of gray matter tissue class; used independently and as an input to the gray-white interface map above. | 10.1016/S1053-8119(09)70884-5 |
| White matter probability | Voxelwise probability map of white matter tissue class; used independently and as an input to the gray-white interface map above. | 10.1016/j.neuroimage.2010.07.033 |

*Gray and white matter probability maps served both as standalone atlases and as inputs to the derived gray-white interface map.*

#### VASCULAR ATLASES *(7 maps)*

| Atlas | Description | Source / DOI |
|---|---|---|
| Cerebral blood flow | Normative map of cerebral blood flow. | 10.1038/s41597-025-05406-w |
| Cerebral blood volume | Normative map of cerebral blood volume, derived from DSC-MRI perfusion imaging. | Fuster-Garcia et al., ISMRM proceedings |
| Arterial structures (TOF-MRA) | Map of large- and medium-sized arterial structures resolvable by time-of-flight magnetic resonance angiography. | 10.3389/fnana.2016.00012 |
| Vessel radius | Normative map of vessel radius, capturing large-to-medium-sized intracranial vasculature. | 10.6084/m9.figshare.c.4215089<br>10.1038/s41597-019-0034-5 |
| Venous density | Normative map of venous density. | 10.1007/s00429-019-01919-4<br>10.6084/m9.figshare.7205960 |
| Venous diameter | Normative map of venous diameter. | 10.1007/s00429-019-01919-4<br>10.6084/m9.figshare.7205960 |
| Venous torsion | Normative map of venous torsion. | 10.1007/s00429-019-01919-4<br>10.6084/m9.figshare.7205960 |

*Vessel radius and arterial structure atlases capture only medium-to-large arterial structures resolvable by MRI and do not reflect microvascular or capillary-level density.*

### METABOLIC ATLASES *(2 maps)*

| Atlas | Description | Source / DOI |
|---|---|---|
| Mitochondrial density | Normative map of mitochondrial density. | 10.1038/s41586-025-08740-6 |
| Glucose metabolism | Normative map of regional glucose metabolism. | 10.1126/sciadv.adi7632 |

### CELL-TYPE ATLASES *(31 maps)*

Spatial maps of cell-type-specific transcriptional programs were derived from the Allen Human Brain Atlas (AHBA) using cell-type marker genes from the Human Brain Cell Atlas (Siletti et al., 2023, Science; marker gene sets defined in Supplemental Table S3 of that publication). For each of the 31 major cell-type superclusters defined by Siletti et al., spatial expression maps of its constituent marker genes were retrieved from the AHBA, and the first principal component of these marker-gene maps was computed to generate a single representative spatial atlas of that cell type's transcriptional program (see Figure 3 for methodological schematic). This procedure was repeated independently for all 31 superclusters, yielding 31 cell-type maps in total.

*Source: 10.1126/science.add7046 (Siletti et al., 2023, Science).*

### GENE EXPRESSION ATLASES *(15633 maps)*

Voxelwise gene expression maps were derived from the Allen Human Brain Atlas (AHBA), a normative atlas of gene expression across the human brain based on post-mortem microarray analysis of six healthy donors sampled across 3,702 brain regions. Maps were generated for all 15,633 genes in the AHBA using abagen (https://github.com/rmarkello/abagen) with recommended preprocessing settings, consistent with published best-practice recommendations for AHBA processing.

Preprocessing steps included: intensity-based filtering (threshold = 0.5), differential stability probe selection, nearest-neighbor interpolation with donor-wise averaging, cross-donor robust sigmoid normalization, and voxel-wise z-scoring. These processed maps are identical to those used in Schaper et al. (2026, DOI pending), and full processing details are described in that publication.

*Sources: 10.1038/nature11405 (Allen Human Brain Atlas); 10.7554/eLife.72129 (abagen); 10.1016/j.neuroimage.2019.01.011 (processing recommendations); Schaper et al., 2026 (DOI to be added when preprint is out)*

### Supplementary Table S3. LAG3 expression across all annotated cell types in the Human Brain Cell Atlas.

*Percentage of cells expressing LAG3, ranked by descending percent expression, across all cell-type annotations in the Human Brain Cell Atlas (Siletti et al., 2023, Science). Cells with >0.50% LAG3 expression are shaded light blue and included in Figure 6C.*

| Cell Type | % LAG3-Expressing Cells | N Cells Expressing LAG3 | N Cells Total |
|---|---|---|---|
| *T cell* | 11.24% | 278 | 2,474 |
| *Purkinje cell* | 7.14% | 3 | 42 |
| *Choroid plexus epithelial cell* | 1.50% | 115 | 7,689 |
| *Natural killer cell* | 1.49% | 10 | 671 |
| *Mammillary body neuron* | 1.30% | 215 | 16,602 |
| *Hippocampal CA4 neuron* | 1.11% | 118 | 10,654 |
| *Thalamic excitatory neuron* | 1.08% | 920 | 85,546 |
| *Cerebellar inhibitory GABAergic interneuron* | 0.90% | 129 | 14,411 |
| *Intratelencephalic-projecting glutamatergic cortical neuron* | 0.76% | 5,170 | 683,473 |
| *Neuron* | 0.54% | 1,972 | 362,316 |
| *Unipolar brush cell* | 0.54% | 5 | 920 |
| *Midbrain-derived inhibitory neuron* | 0.50% | 637 | 126,782 |
| *Hippocampal CA1-3 neuron* | 0.41% | 311 | 74,979 |
| *Oligodendrocyte precursor cell* | 0.38% | 401 | 105,734 |
| *Cerebellar granule cell* | 0.33% | 449 | 135,835 |
| *Medial ganglionic eminence derived interneuron* | 0.30% | 677 | 222,434 |
| *Amygdala excitatory neuron* | 0.30% | 333 | 109,452 |
| *Pvalb chandelier GABAergic interneuron* | 0.29% | 42 | 14,486 |
| *Sst chodl GABAergic interneuron* | 0.29% | 11 | 3,850 |
| *Vascular associated smooth muscle cell* | 0.28% | 3 | 1,074 |
| *Cerebral cortex neuron* | 0.27% | 214 | 78,396 |
| *Near-projecting glutamatergic cortical neuron* | 0.24% | 45 | 18,856 |
| *Lamp5 Lhx6 neuron* | 0.23% | 69 | 30,632 |
| *Caudal ganglionic eminence derived interneuron* | 0.22% | 501 | 227,671 |
| *Central nervous system macrophage* | 0.21% | 7 | 3,344 |
| *Medium spiny neuron* | 0.20% | 297 | 152,189 |
| *Eccentric medium spiny neuron* | 0.19% | 75 | 40,144 |
| *Dentate gyrus granule cell* | 0.18% | 124 | 67,533 |
| *Pericyte* | 0.16% | 6 | 3,693 |
| *Fibroblast* | 0.15% | 14 | 9,156 |

| Cell Type | % LAG3-Expressing Cells | N Cells Expressing LAG3 | N Cells Total |
|---|---|---|---|
| *Microglial cell* | 0.10% | 92 | 88,494 |
| *Ependymal cell* | 0.10% | 6 | 5,882 |
| *Endothelial cell* | 0.10% | 5 | 5,165 |
| *Astrocyte* | 0.07% | 104 | 155,025 |
| *Committed oligodendrocyte precursor* | 0.04% | 2 | 4,720 |
| *Bergmann glial cell* | 0.02% | 2 | 8,041 |
| *Oligodendrocyte* | 0.02% | 83 | 490,246 |
| *Monocyte* | 0.00% | 0 | 503 |
| *B cell* | 0.00% | 0 | 105 |
| **Total (all cell types)** | **0.40%** | **13,445** | **3,369,219** |
